# Genomic profiling reveals heterogeneous populations of ductal carcinoma in situ of the breast

**DOI:** 10.1101/2020.06.22.20137018

**Authors:** Satoi Nagasawa, Yuta Kuze, Ichiro Maeda, Yasuyuki Kojima, Ai Motoyoshi, Tatsuya Onishi, Tsuguo Iwatani, Takamichi Yokoe, Junki Koike, Motohiro Chosokabe, Manabu Kubota, Hibiki Seino, Ayako Suzuki, Masahide Seki, Katsuya Tsuchihara, Eisuke Inoue, Koichiro Tsugawa, Tomohiko Ohta, Yutaka Suzuki

**Author notes:** Correspondence to: Dr. Yutaka Suzuki, Department of Computational Biology and Medical Sciences, Graduate School of Frontier Sciences, The University of Tokyo, 5- 1-5, Kashiwanoha, Kashiwa-shi, Chiba 277 8561, Japan. Equally contributed author.

## Abstract

A substantial number of cases of ductal carcinoma in situ (DCIS) of the breast will never progress to invasive ductal carcinoma (IDC), indicating they are overtreated under the current criteria. Although various candidate markers are available, the relevant markers for delineating the risk categories have not been established. In this study, we analyzed of the integrated clinical features of 431 cases of DCIS followed by deep sequence analyses in a 21-case discovery cohort and a 72-case validation cohort. We identified the five most critical markers of the aggressiveness of DCIS: age <45 years, *HER2* amplification, *GATA3* mutation positivity, *PIK3CA* mutation negativity, and PgR protein negativity. Spatial transcriptome and single-cell DNA sequencing further revealed that *GATA3* dysfunction, but not *PIK3CA* mutation, upregulates EMT, invasion, and angiogenic pathways followed by PgR downregulation. These results reveal the existence of heterogeneous populations of DCIS and provide predictive markers for classifying DCIS and optimizing treatment.

## Introduction

Non-invasive ductal carcinoma of the breast, i.e., ductal carcinoma in situ (DCIS), is an early stage of breast cancer that may potentially develop into invasive ductal carcinoma (IDC)^1,2^. Because the breast duct is anatomically free of blood or lymphatic vessels, the lesions localized in situ theoretically do not undergo metastasis. The incidence of DCIS has been increasing because of improvements in diagnostic modalities, permitting preventive surgery based on the assumption that surgical excision reduces the risk of eventual IDC development^3–5^. However, it has been noted that a substantial number of DCIS lesions diagnosed using the current criteria may not progress to IDC in the absence of treatment^6–11^. Clinically, DCIS can be divided into three categories: 1) unlikely to progress to IDC even without surgical treatment (low-risk or false DCIS); 2) truly precancerous lesions of IDC (true DCIS); 3) high potential for recurrence as IDC even with standard treatment (high-risk DCIS). It is critical to discriminate low-risk DCIS from true DCIS to avoid unnecessary surgery, thereby improving patient quality of life and reducing medical costs. Contrarily, intensive treatment should be provided for patients with high-risk DCIS to improve their prognosis. Therefore, an accurate classification for directing treatment is an urgent clinical challenge. Indeed, global clinical trials are underway in several countries to determine whether non-resection treatment is feasible for low-risk DCIS^12–14^. However, the current three-category classification system is not straightforward. The current classification depends on clinicopathological factors including age, tumor size, presence of comedo necrosis, nuclear grade, and the hormone receptor and human epidermal growth factor receptor 2 (HER2) status^15–19^, and the evaluations vary among pathologists or institutes in the absence of a global standard^20^. One critical issue that has not been resolved is the lack of biological verification of the risk factors that contribute to the malignant transformation of DCIS *in vivo*. The small amounts of tumor specimens available for analysis and the heterogeneity of tumors have made such studies of DCIS difficult. To address this issue, we identified objective clinicopathological and genomic risk factors for DCIS recurrence based on an analysis of the integrated clinical features of 431 patients with pure DCIS, which lacks any trait of IDC, followed by whole-exome analysis in a discovery cohort of 21 patients with DCIS and targeted deep sequence analysis in a validation cohort of 72 patients with DCIS. Then, we examined the contributions of these to the progression of invasive cancer *in vivo* using spatial transcriptome sequencing (STseq) and single-cell DNA sequencing (scDNA-seq). STseq is a novel technique that provides gene expression information on pathological sections^21,22^. It allows gene expression profiling without losing the positional information about the different cell types that constitute tumors *in vivo*. In this study, the combined sequencing techniques allowed us to clarify the significance of risk stratification factors and their biological consequences *in vivo* for the first time.

## Results

### Clinicopathological risk factor determination

The records of 431 patients with pure DCIS who underwent surgery at the St. Marianna University School of Medicine from 2007 to 2012 were reviewed to determine the clinical criteria for stratifying patients with low- or high-risk DCIS. The median age of patients at diagnosis was 48 years (range, 24–90 years). The median follow-up period was 6.1 years (range, 0.5–10.9 years). Twenty lesions (4.6%) progressed to IDC during the follow-up period. Three hundred seventy-five lesions (87%) were positive for estrogen receptor (ER) expression. Eighty-one lesions (18.8%) were positive for *HER2* amplification. The detailed clinicopathological data are presented in SourceData1. Univariate analysis using Cox proportional hazards regression models was performed to assess the relationship between predictive factors and relapse-free survival (Table 1). Age (≥45 years vs. <45 years, determined via receiver operating curve [ROC] curve analysis, area under the curve [AUC] = 0.67, Supplementary Fig. S1) and *HER2* amplification status appeared to be significantly and independently associated with relapse in multivariate analysis, with hazard ratios (HRs) of 3.86 (95% confidence interval [CI] = 1.53–9.73, P = 0.00425) and 3.45 (95% CI = 1.34–8.5, P = 0.00717), respectively. Notably, the HR for *HER2* positivity and age <45 years (vs. *HER2* negativity and age ≥ 45 years) was 11.03 (95% CI = 3.54–34.4, P < 0.0001). Based on these results, we decided to use age and the *HER2* amplification status as the major clinical criteria for discriminating low- and high-risk patients in the subsequent analyses. The presence/absence of comedo necrosis and nuclear atypia, which are traditional risk evaluation criteria, were excluded and only used for reference purposes.

### Selection of genomic risk factors in whole-exome sequencing

To improve the accuracy of the criteria for discriminating low- and high-risk DCIS based on the clinical evaluation, we attempted to identify genomic factors. As a discovery cohort, we analyzed 21 patients with pure DCIS who had been randomly selected with a matched intrinsic type and relatively high recurrence rate. The clinicopathological characteristics of patients in the discovery cohort are shown in Supplementary Table. S1. Whole-exome libraries were constructed from tissue samples obtained via microdissection (Supplementary Fig. S2) and subjected to sequencing analysis at a sufficient sequencing depth (averaging ×207, SourceData2). Matched normal breast tissue was sequenced (average depth, ×109.1) in parallel to distinguish germline variants from somatic mutations. The obtained mutation graph is shown in Fig. 1a.

**Fig. 1.**
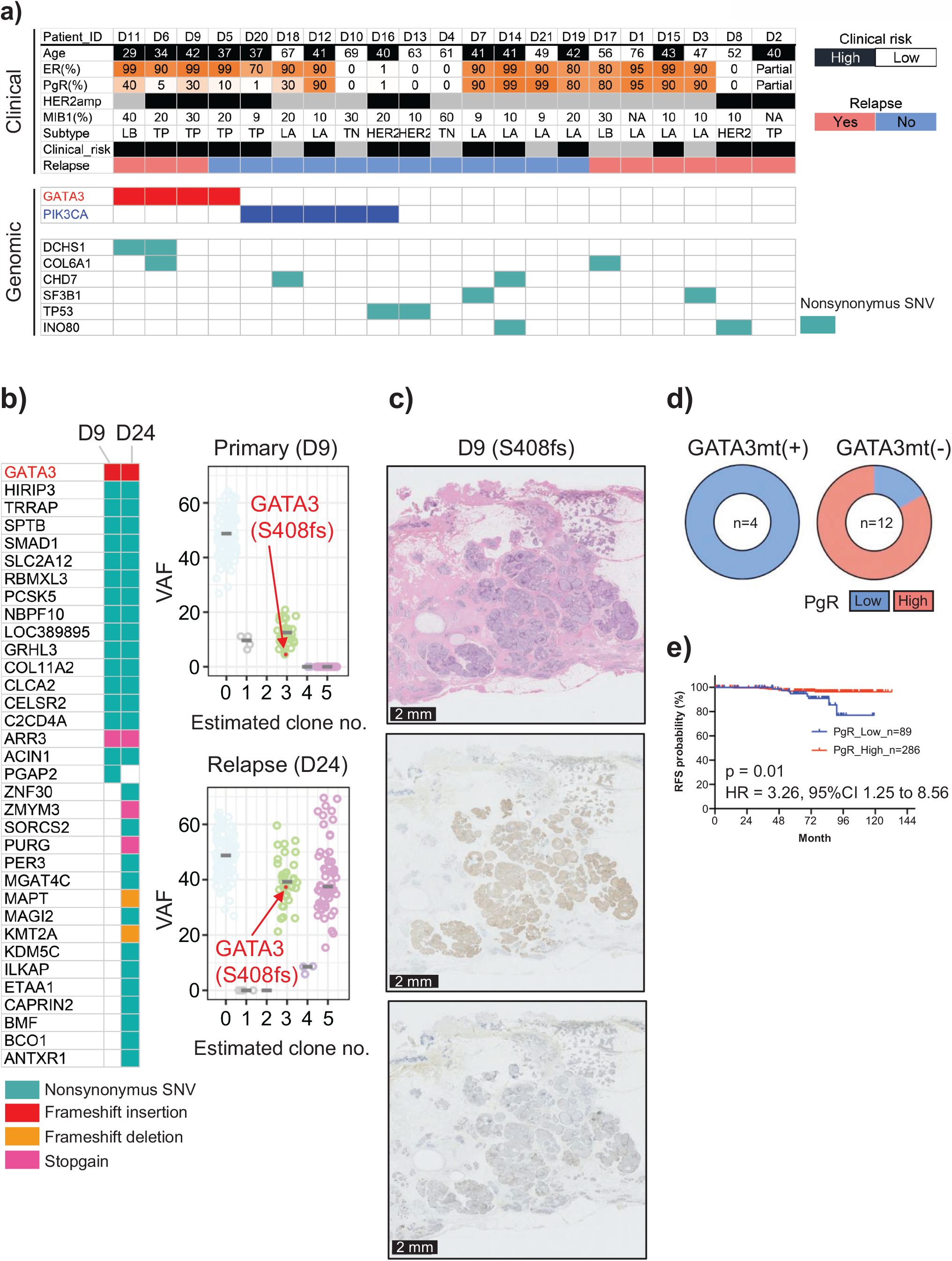
Selection of clinicopathological and genomic risk factors for recurrence. **1a)** Results of whole-exome sequencing of 21 patients with primary pure ductal carcinoma in situ (DCIS). **Upper:** The clinical information included age, the percentages of estrogen receptor (ER)-, progesterone receptor (PgR)-, and MIB1-positive cells by immunohistochemistry (IHC), human epidermal growth factor receptor 2 (*HER2*) expression status, and recurrence status. Patients were subdivided into molecular phenotypes using IHC surrogates (luminal A [ER−/PR+/HER2−], luminal B [ER+/PR+/HER2+ or ER+/PR+/HER2−/MIB1 index high], HER2 [ER−/PR−/HER2+], and TN [ER−/PR−/HER2−]). Clinical risk estimated by age and *HER2* expression status basis on the results in Table 1 is shown. **Middle:** *GATA3* and *PIK3CA* mutations are shown (red and blue, respectively). Only the results for non-synonymous mutations are shown. In total, 4 of 21 (19%) patients carried non-synonymous *GATA3* mutations. Non-synonymous *PIK3CA* mutations were detected in 5 of 21 (24%) patients. **Lower:** The genes that overlapped among patients are shown. Only the results for non-synonymous mutations are shown. **1b)** Comparison of *GATA3* mutations between primary and relapse tumors. **Left:** Comparison of mutations via whole-exome sequencing of the primary (D9) and matched relapse lesions (D24) are shown. The same *GATA3* mutation (S408fs) was detected in both lesions. **Right:** Results of subclone analysis obtained using PyClone software are shown. The findings suggest that the subclone carrying the *GATA3* mutation also existed in the relapse lesion. VAF: variant allele frequency. **1c)** Panel shows HE (upper panel), ER (middle panel), and PgR (lower panel) staining for patient D11 with a *GATA3* frameshift mutation (S408fs). Although ER expression was positive, PgR expression was decreased. Scale bars: 2 mm. HE: hematoxylin and eosin; VAF: variant allele frequency. **1d)** Pie charts depicting PgR expression was assessed via IHC in 16 ER-positive cases with and without *GATA3* mutations. IHC revealed that in patients with ER-positive DCIS, *GATA3* mutation was significantly associated with reduced PgR expression (P = 0.0192 by Fisher’s exact test). **1e)** Kaplan–Meier curve for 375 patients with ER-positive DCIS according to PgR expression. High PgR expression was indicated by positive immunostaining in ≥70% of cells, and low expression was indicated by positive staining in ≤60% of cells. The prognosis of ER-positive patients with low PgR expression was significantly worse than that of patients with high PgR expression (P = 0.01 by log-rank test, hazard ratio = 3.26, 95% confidence interval = 1.25–8.56).

*GATA3* and *PIK3CA* mutations were the most commonly detected mutations in the cohort, being present in four (19%) and five patients (24%), respectively. Of the four patients with *GATA3* mutation, three experienced recurrence of IDC. The odds ratio (OR) for relapse for *GATA3* mutation positivity versus *GATA3* mutation negativity was 5.5 (95% CI = 0.63–76.7). To further investigate the significance of *GATA3* mutation to relapse, samples from patients with recurrent tumors were additionally analyzed for mutation status in comparison with their paired primary DCIS lesions. Importantly, two of the four patients harbored the same *GATA3* mutation in their primary and recurrent tumors (Fig. 1b, Supplementary Fig. S3), suggesting that relapse was caused by a *GATA3*-mutated clone from the primary lesions. This finding was supported by the results of computational modeling performed to detect cancer evolution using the PyClone program^23^. From these results, we identified *GATA3* mutation as a candidate genomic high-risk factor in the discovery cohort. It was recently reported that *GATA3* variants with altered DNA-binding capacity led to reduced expression of the gene set containing the progesterone receptor (*PgR*) gene and increased expression of genes involved in epithelial-to-mesenchymal transition (EMT), such as cell movement and cell invasion pathways^24^. We therefore examined PgR protein expression via immunohistochemical analysis in 16 ER-positive patients including the four patients who harbored *GATA3* mutations. PgR expression was decreased in all four patients with *GATA3*-mutated DCIS, whereas only two of the remaining 12 patients exhibited reduced PgR expression (Fig. 1c and 1d, Supplementary Fig. S4). We found that *GATA3* mutation was associated with reduced PgR expression (Supplementary Table S2, P = 0.0192 by Fisher’s exact test). To investigate the association between prognosis and PgR expression in patients with ER-positive DCIS, we examined 375 patients with ER-positive DCIS among the clinical cohort of 431 patients. PgR expression was determined using ROC curve analysis, revealing an AUC of 0.66 (Supplementary Fig. S5). ER-positive patients with low PgR expression (≤60%) displayed a significantly high incidence of relapse than those with high PgR expression (Fig. 1e; p = 0.01, HR = 3.26, 95% CI = 1.25– 8.56). Together, low PgR expression, which is routinely measured in clinical practice, likely represents a high-risk factor that could be a surrogate marker for *GATA3* mutation in patients with ER-positive DCIS.

Contrary to the findings for *GATA3* mutation, no patient with *PIK3CA* mutation experienced recurrence, whereas 9 of 16 patients without *PIK3CA* mutation (56.2%) experienced recurrence. Although *PIK3CA* mutation is a known cancer driver in many types of cancer^25^, interestingly, it has been associated with better prognosis in invasive breast cancer compared with wild-type *PIK3CA*^26^. Thus, we identified *PIK3CA* mutation as a candidate genomic low-risk factor for DCIS in the discovery cohort.

### Validation analysis of the risk factors via targeted deep sequencing

To validate the findings revealed using the clinical data set and discovery cohort, we conducted an analysis using a larger and independent validation cohort of 72 patients. The clinicopathological characteristics of these patients are shown in Supplementary Table. S1. To expedite the sequencing, a custom sequencing panel was designed on the basis of the observed mutation spectrum of the discovery cohort, which included 180 genes (SourceData3). The median follow-up period was 5.5 years (range, 0.5– 9.6 years). Of 72 patients, nine experienced relapse. In this cohort, high clinical risk as determined using the previously identified criteria (Table 1) tended to be correlated with poor outcome, albeit without statistical significance (OR = 1.1, 95% CI = 0.25– 4.5).

The target sequencing was conducted at a sufficient sequencing depth (averaging ×592.2). The obtained mutation graph is shown in Fig. 2. In total, 40 (56%) and 36 patients (50%) harbored *GATA3* and *PIK3CA* mutations, respectively. In concordance with the discovery cohort results, *GATA3* mutation was positively associated with relapse (OR = 7.8; 95% CI = 1.17–88.4), whereas *PIK3CA* mutation was negatively associated with relapse (OR = 0.45; 95% CI = 0.12–1.7). Of the 180 genes tested, none was superior to these two factors as a predictive marker. Concerning PgR protein expression, low PgR expression was again correlated with relapse in the validation cohort (OR = 25.6; 95% CI = 3.64–142.2). Taking these findings into consideration, we believe that the results from the discovery cohort reflect the true nature of DCIS.

**Fig. 2.**
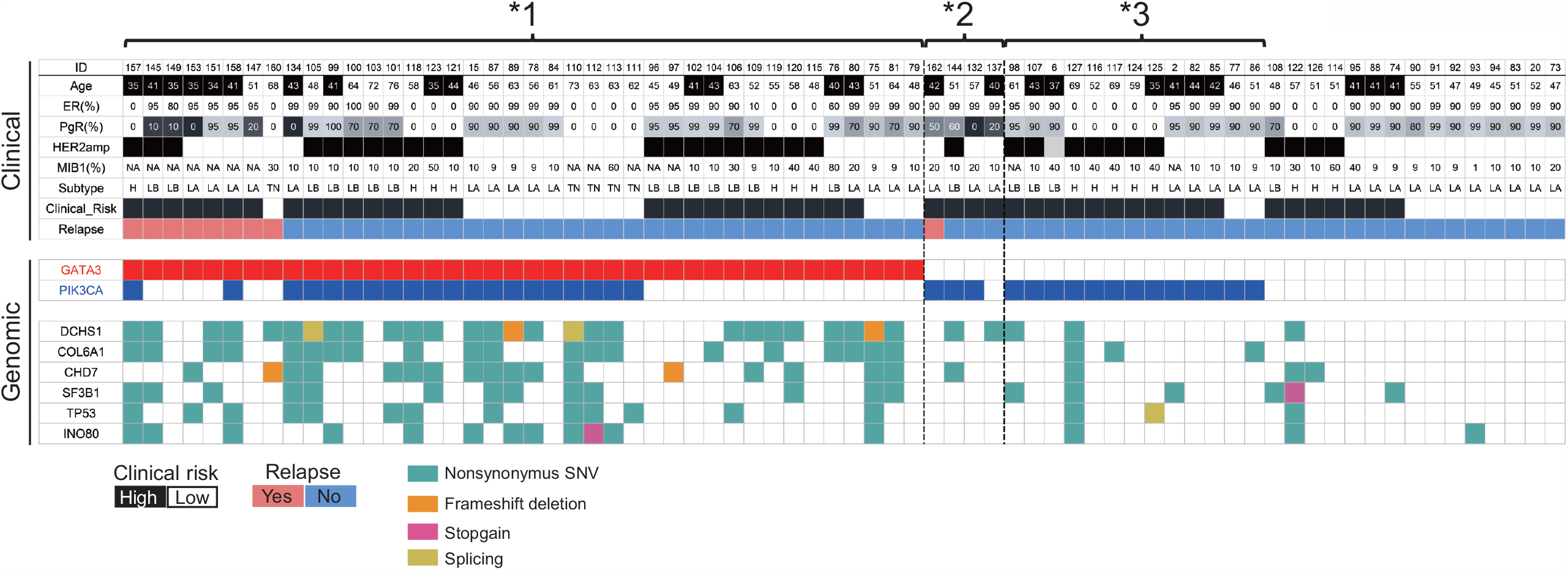
Validation of clinicopathological and genomic risk factors via targeted deep sequencing. **Upper:** The clinical information included age, percentages of estrogen receptor (ER)-, progesterone receptor (PgR)-, and MIB1-positive cells according to immunohistochemistry (IHC), human epidermal growth factor receptor 2 (*HER2*) expression status, and recurrence status. Patients were subdivided into molecular phenotypes using IHC surrogates (luminal A [ER−/PR+/HER2−], luminal B [ER+/PR+/HER2+ or ER+/PR+/HER2−/MIB1 index high], HER2 [in figure, shown as H; ER−/PR−/HER2+], and TN [ER−/PR−/HER2−]). Clinical risk estimated by age and *HER2* expression status basis is shown. **Middle:** *GATA3* and *PIK3CA* mutations are shown (red and blue, respectively). Only the results for non-synonymous mutations are presented. In total, 40 of 72 (56%) patients carried non-synonymous *GATA3* mutations. Non-synonymous *PIK3CA* mutations were detected in 36 of 72 (50%) patients. **Lowe**r: The genes that overlapped among patients in the discovery cohort are shown. *1: *GATA3* mutation-positive ductal carcinoma in situ (DCIS). *2: *GATA3* mutation-negative, ER-positive DCIS with low PgR expression. *3: *PIK3CA* mutation-positive DCIS that does not satisfy *1 or *2.

### Analyses of DCIS cases for molecular dissection

To dissect the molecular etiology underlying the identified high-risk markers, we precisely investigated fresh frozen specimens from three additional representative patients. In particular, according to STseq and scDNA-seq, Case A had DCIS with a *GATA3* mutation (Fig. 2, *1) and a microinvasion, and the lesion was regarded as true DCIS with progression. Case B had DCIS with low PgR expression (Fig. 2, *2) and a microinvasion, and the lesion was regarded as true DCIS. Case C had DCIS with a *PIK3CA* mutation (Fig. 2, *3) without any microinvasion, and the lesion was regarded as possibly false DCIS (low-risk DCIS). The clinicopathological information of each case is provided in Supplementary Table S3.

### *STseq of a DCIS lesion harboring a* GATA3 *mutation*

First, we profiled the spatial gene expression of the specimen from Case A via STseq using the Visium platform of 10x Genomics (Pleasanton, CA, USA), which is a recently developed barcoding-based spatial transcriptomics technology. This patient was diagnosed with DCIS in the preoperative pathological diagnosis but found to have a site of microinvasion in the postoperative pathological diagnosis. Thus, this DCIS lesion was identified as a true precursor for IDC. The panel sequencing analysis at an average depth of ×2391.4 revealed this sample harbored a *GATA3* mutation (exon4:c.865dupG:p.C288fs, variant allele frequency [VAF] = 7%). Interestingly, a *PIK3CA* mutation was also detected at a higher VAF (exon5:c.T1035A:p.N345K, VAF = 26.1%), suggesting that the emerging *GATA3* mutation is overwriting the basal features of DCIS with the *PIK3CA* mutation. The full mutation list is provided in SourceData4.

For the Visium analysis, we collected 799,133,428 sequencing reads at a sequencing saturation of 86.2%. The number of analyzed spots (at 55 µm in diameter) was 2043, which contained a median of 7469 UMIs or a median of 2928 detected genes per spot (SourceData5). As shown in the middle panel of Fig. 3a, cancer cell spots were classified into three groups via un-hierarchical k-means clustering (k = 9), suggesting the presence of heterogeneity in the DCIS lesion. Additionally, non-cancer spots, which may represent the microenvironment surrounding the cancer cells, were classified into four clusters. Among a total of 422 spots representing pathologically identified cancer cells, *GATA3* mutation reads were detected in 46 spots by analyzing Visium reads (Fig. 3b, Supplementary Fig. S6). Fortunately, in this case, the *GATA3* mutation was located at the 3’-end of the transcript, and thus could be represented by the Visium reads.

**Fig. 3.**
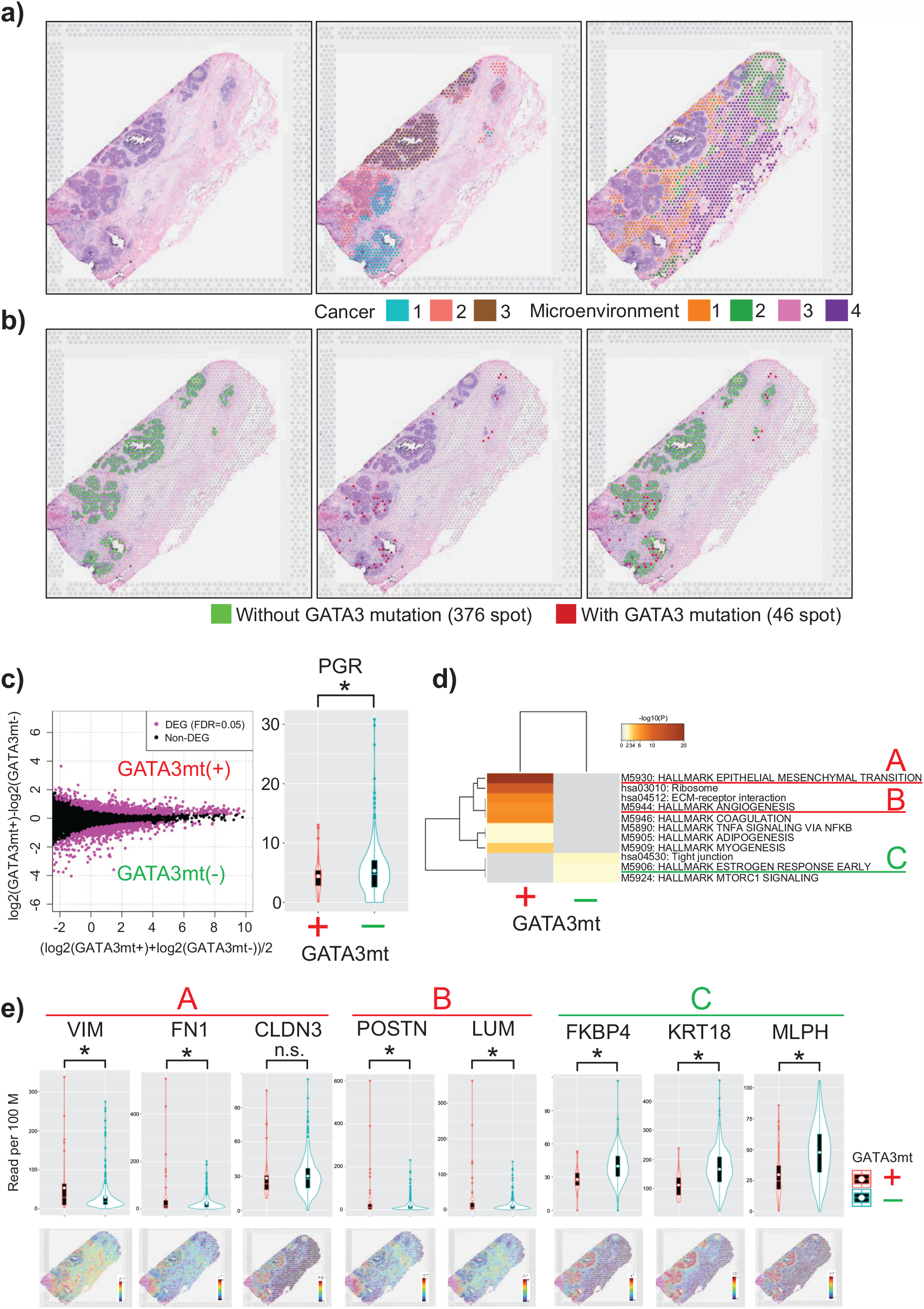
Spatial transcriptome analysis of a patient with *GATA3* mutation-positive ductal carcinoma in situ (DCIS) (Case A) **3a)** Visualization of the Visium results of Case A. Hematoxylin and eosin staining (left). In total, 2043 spots in the tissue are represented. Morphologically, those cells were classified as cancer cells in situ (405 spots) and non-cancer cells (1638 spots). Three cancer cell populations as classified via un-hierarchical k-means clustering (k = 9) are shown with different colors (middle panel). Four non-cancer spots are similarly shown to represent the gene expression reflecting the microenvironments at the corresponding positions (right panel). **3b)** The spots in which *GATA3* mutations were detected as transcriptomic tags are colored red. Of the 422 spots morphologically located with cancer cells, a *GATA3* mutation read was found for 46 spots. **3c)** MA plot presenting differentially expressed genes (DEGs) between spots with and without *GATA3* mutations. In total, 1468 DEGs were detected at a false discovery rate of <0.05 (left). The right panel shows a comparison of progesterone receptor (*PgR*) expression at each spot. Decreased *PgR* expression was more frequently observed in the spots with *GATA3* mutations (P = 0.02901, one-sided *t*-test). **3d)** Results of the enrichment test for the MSigDB v5.1 Hallmarks gene set collection (HALLMARK) and Kyoto Encyclopedia of Genes and Genomes (KEGG) pathways are presented. Each band represents one enriched term or pathway. The level of enrichment is color-coded as the −log 10 p value. As indicated, in the spots with *GATA3* mutations, the DEGs were mainly enriched in epithelial-mesenchymal transition (“A”) and angiogenesis (“B”). Conversely, in the spots without *GATA3* mutations, the DEGs were mainly enriched in tight junctions, estrogen response (“C”), and MTORC1 signaling. **3e)** Violin plots showing the expression of the representative genes corresponding to the gene groups of A–C. The expression of the indicated gene was compared between the spots with and without *GATA3* mutations in the upper panel. * P < 0.05 (one-sided), n.s.: not significant. The gene expression levels in the tissue are shown in the bottom panel.

Using TCC R package^27^, we detected differentially expressed genes (DEGs) between the spots with and without *GATA3* mutations. In total, 1468 DEGs were detected (false discovery rate [FDR] < 0.05, Fig. 3c, left panel). As identified in Fig. 1d, decreased PgR expression was observed in the spots with *GATA3* mutations (p = 0.02901; *t*-test; Fig. 3c, right panel). To predict the functional consequence of the detected DEGs, we conducted pathways analysis using Metascape (http://metascape.org/)^28^. In the cancer spots with *GATA3* mutations, nine key pathways, which were associated with 11 cancer hallmarks, were affected compared with the findings in the mutation-negative spots (Fig. 3d). In particular, the key genes and pivotal pathways included EMT and angiogenesis. Importantly, the key genes with expression changes were those identified to occur in response to aberrant *GATA3* function, such as *VIM* and *FN1* (Fig. 3e). These results indicate that *GATA3* mutation arises during DCIS progression accompanied by malignant features. Because it has been reported that increased VIM expression occurs downstream of *GATA3* mutation in luminal cells^24^, the observed *GATA3* mutations in the spots could represent prior genetic alterations during malignant development. Conversely, in the spots without *GATA3* mutations, the DEGs were mainly enriched in estrogen response, tight junction, and mTORC1 signaling pathways, suggesting that the cells in those spots acquired the minimum changes permitting cell transformation.

### STseq of a DCIS lesion with low PgR expression but no GATA3 mutation

We next investigated Case B as a rather exceptional case with low PgR expression despite no *GATA3* mutation (Fig. 2, *2). From the target sequencing at an average of depth of ×3030.6, a *PIK3CA* mutation was detected (exon21:c.A3140T:p.H1047L, VAF:40%), whereas no *GATA3* mutation was detected even at this sufficient sequencing depth. Therefore, this case was expected to have a good prognosis on a molecular basis. Nevertheless, postoperative pathological examination revealed a microinvasion, indicating that this lesion was a true precursor of IDC. Immunohistochemical analysis illustrated that the lesion was ER-positive and PgR-negative.

The Visium analysis uncovered heterogeneous gene expression depending on the location of cells for both cancerous and non-cancerous regions (Supplementary Fig. S7). PgR expression was reduced despite the observed high ER expression (Fig. 4a, upper panel), consistent with the immunostaining findings (Fig. 4a, lower panel). Focusing on the gene expression changes in DCIS cells, 188 spots that were morphologically located in the intraductal regions were manually selected. Unsupervised hierarchical clustering of these spots identified three apparent clusters (Fig. 4b, left panel). Spatially, the different clusters corresponded to different regions (Fig. 4b, middle panel). Importantly, the 48 spots of cluster 1 (colored red) overlapped with the location of DCIS cells that were about to invade on the basis of their morphology (indicated by the red arrow head in the right panel of Fig. 4b). Meanwhile, the 101 spots of cluster 2 (colored green) were located in the center of the same duct, and they were regarded as cells that were not invading the stroma anatomically (indicated by the green spots in the middle panel of Fig. 4b). Differential expression analysis between clusters 1 and 2 identified 2747 DEGs (FDR < 0.05, Fig. 4c, left). Interestingly, *GATA3* expression was significantly decreased in cluster 1 (Fig. 4c, right). It is likely that functional aberration of *GATA3* was invoked at the gene level in this patient despite the absence of a mutation of this gene. Consistently, *GATA3*-centered gene expression changes were observed in this patient as judged by the results of gene enrichment and pathway analysis, as observed for Case A (Fig. 4d, 4e). A total of 21 pathways and 18 cancer hallmarks, including EMT and angiogenesis pathways, were affected. As observed for Case A, in cluster 2, which had not invaded the stroma, the DEGs mainly represented estrogen response-related genes. Taken together, we concluded that, in this case too, *GATA3* plays pivotal roles via its expression changes. Even without the genomic mutations, aberrant expression of GATA3 may result in an equivalent consequence in some patients.

**Fig. 4.**
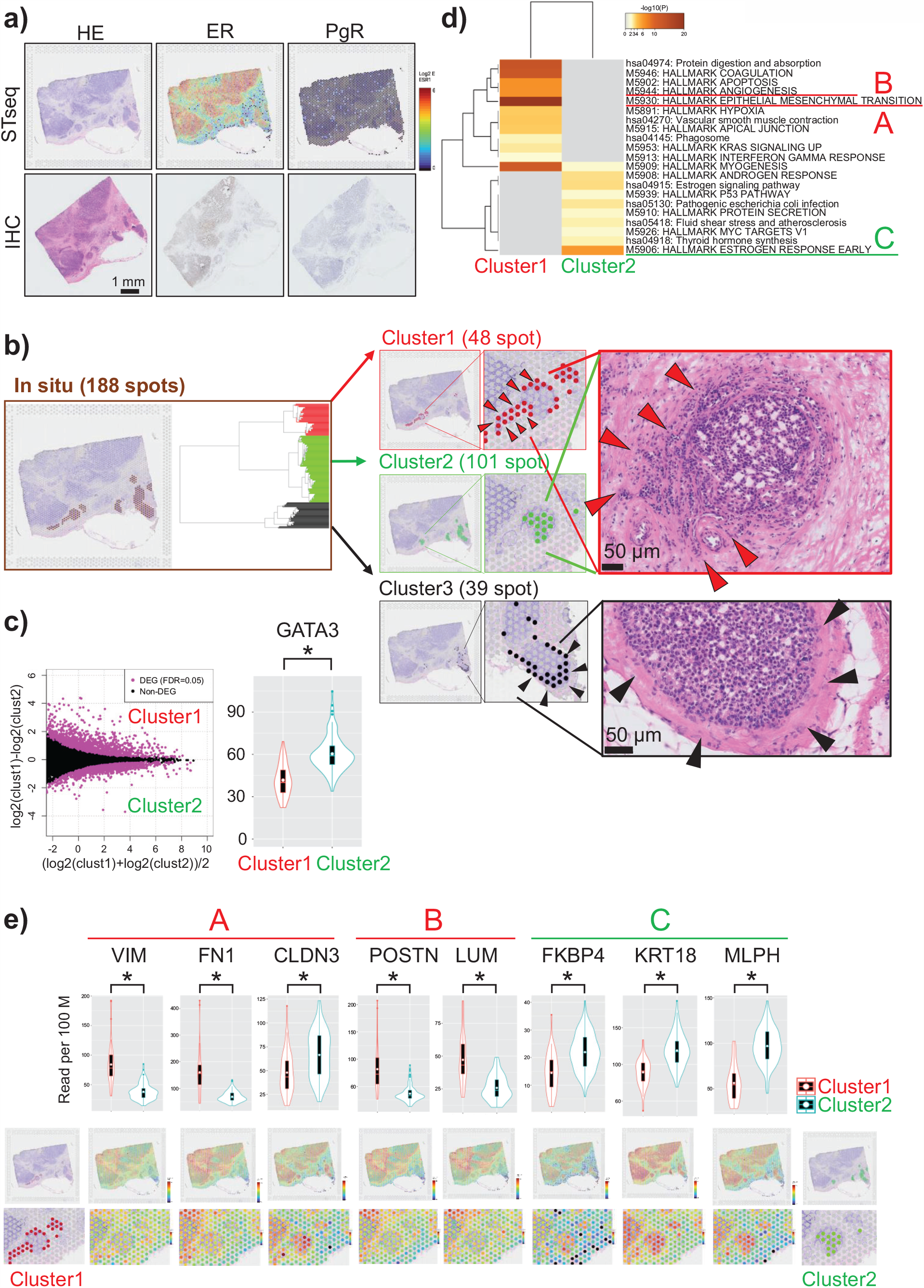
Spatial transcriptome analysis of low progesterone receptor (PgR) expression in a patient with estrogen receptor (ER)-positive ductal carcinoma in situ (DCIS) (Case B) **4a)** Visualization of the Visium results of Case B. The upper row shows the expression of genes indicated in the top margin. The lower panels show the results of hematoxylin and eosin staining and immunostaining for ER and PgR. **4b)** Dendrogram generated from the hierarchical clustering performed across 188 spots indicated as colored dots (left panel). The middle panel presents the three distinct clusters detected via hierarchical clustering. The right panel shows the originating positions of the indicated spots in the tissue. The rightmost panels present the following observations. Namely, the 48 spots of cluster 1 overlapped with the locations of DCIS cells that were about to invade (red arrow) on the basis of their morphology. The spots of cluster 2 were located in ducts that were anatomically in the same position as those of cluster 1, but their locations coincided with those of DCIS cells that had not invaded. The location of cluster 3 was consistent with that of DCIS cells that were not invasive (black arrow) on the basis of their morphology. **4c)** MA plot showing differentially expressed genes (DEGs) between clusters 1 and 2. In total, 2747 DEGs was detected (false discovery rate < 0.05, left). The right panel shows a comparison of *GATA3* expression (P < 0.001, one-sided *t*-test). Decreased *GATA3* expression was observed in cluster 1. **4d)** MSigDB v5.1 Hallmarks gene set collection (HALLMARK) and Kyoto Encyclopedia of Genes and Genomes (KEGG) pathway analyses were conducted using the selected DEGs (top 100 genes were used) and the Metascape tool. Each band represents one enriched term or pathway colored according to the −log 10 p value. In spots in cluster 1, the DEGs were mainly enriched in epithelial-mesenchymal transition (“A”) and angiogenesis (“B”). Conversely, in spots in cluster 2, the DEGs were mainly enriched in estrogen response (“C”). **4e)** Violin plots showing the expression of each gene in A, B, and C in spots from clusters 1 and 2 (upper panel). Visualization of the expression of each gene on the Visium slide (lower panel). * P < 0.05 (one-sided), n.s.: not significant.

### *Integrated STseq and scDNA-seq analysis of a patient with DCIS and* PIK3CA *mutation*

Lastly, we analyzed the lesion of Case C in the discovery cohort as a possibly false DCIS that was unlikely to progress to IDC. This case belonged to the group indicated as *3 in Fig. 2 in the validation cohort, which was typified by *PIK3CA* mutation, the absence of *GATA3* mutation or downregulation, no microinvasion, and no relapse after surgery. The DCIS lesion in this case harbored two mutations in *PIK3CA* (exon10:c.G1633A:p.E545K, exon10:c.A1634G:p.E545G, VAF = 25% for both mutations) as determined via whole-exome sequencing with an average depth of ×134.8.

The Visium analysis revealed a monotonous gene expression pattern, in line with the morphologically monoclonal structure of this cancer (Fig. 5a, left panel). Indeed, the spots were roughly separated into two clusters via un-hierarchical clustering (Fig. 5a, middle and right panels). They almost completely overlapped with the morphologically determined cancer and non-cancer cells (stromal cells). Thus, we compared 347 spots in the cancer cells (blue spots) with 151 spots in the stromal cells (orange spots) using DEG analysis and identified 508 upregulated DEGs in the cancer spots (FDR < 0.05). Weighted gene co-expression network analysis (WGCNA) revealed that only two major nodes (modules) were disturbed in the cancer spots compared with the non-cancer spots (Fig. 5b, upper panel). We conducted subnetwork analysis to identify the hub genes (Fig. 5b, middle and lower panels). Pathway enrichment analysis of the hub genes revealed that the genes involved in estrogen response and p53 pathways were enriched in module 1 (Fig. 5c), which shares the characteristics observed for the benign-appearing spots in Cases A and B, indicating that malignant transition had not occurred in this lesion.

**Fig. 5.**
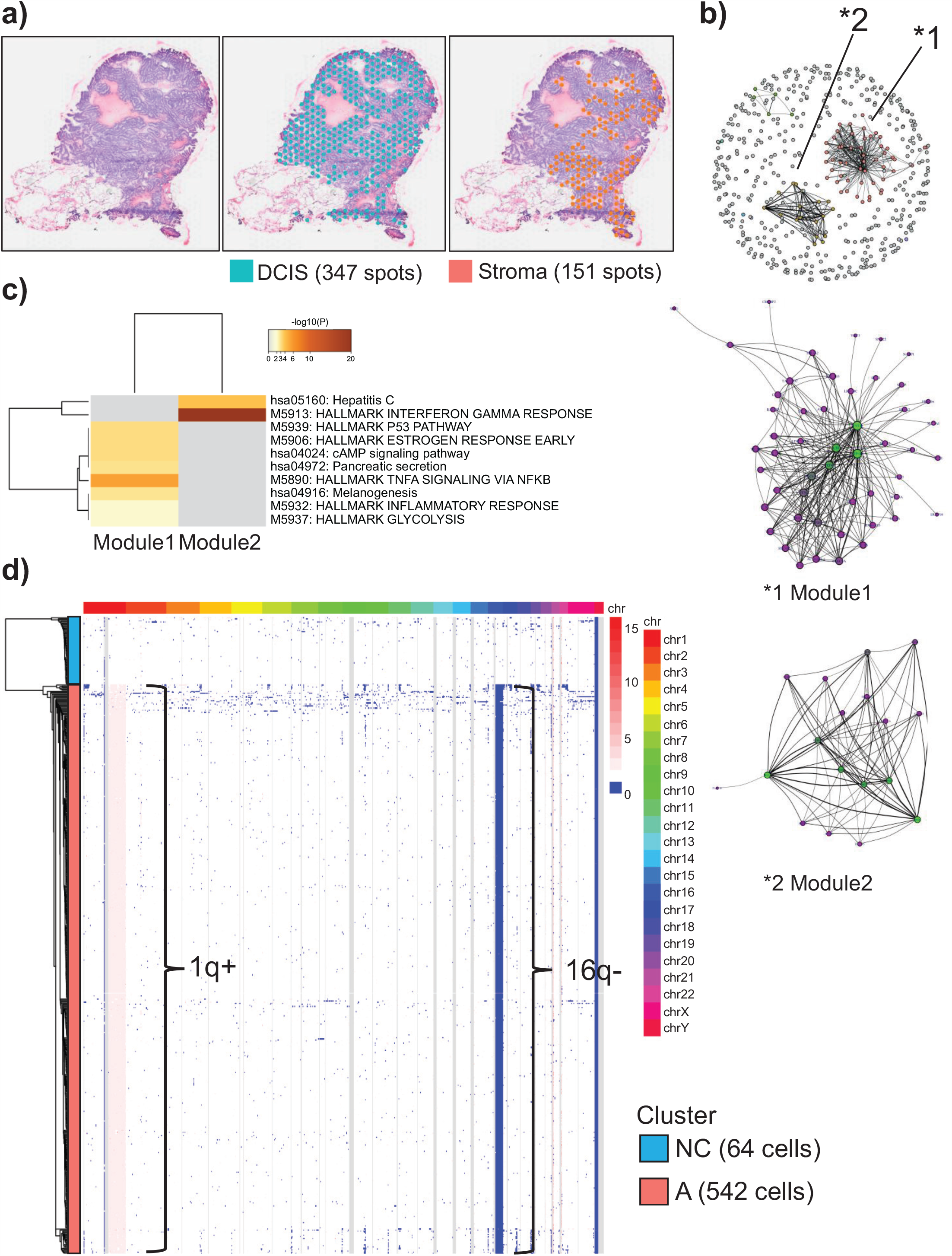
Integrated spatial transcriptome and single-cell DNA sequencing analysis of a patient with *PIK3CA* mutation-positive ductal carcinoma in situ (DCIS) 5a) Visualization of the Visium results of Case C. Hematoxylin and eosin staining (left). The number of spots in the tissue was 594. Spots were classified into two clusters via un-hierarchical k-means clustering. They clusters almost completely overlapped with morphologically identified cancer cells (middle panel) and non-cancer cells (stromal cells, right panel). **5b)** Co-expression network analysis of transcriptomes in DCIS spots. Weighted gene co-expression network analysis was applied to build the co-expression network and identify gene modules. The nodes and edges indicate genes and significant correlations between genes, respectively. In the top panel, the two nodes that appeared to have changed from normal cells are indicated by asterisks. Middle and lower panels present the subnetworks in modules (*1) and (*2), respectively. Node centrality, defined as the sum of strength, is represented as the node size and its color. **5c)** Pathway analysis was performed for each module using the Metascape tool. Each band represents one enriched term or pathway colored according to the −log 10 p value. Pathway enrichment analysis revealed that genes involved in p53 and estrogen signaling pathways were enriched in module 1, which shares the characteristics observed for the benign-appearing spots in Cases A and B, indicating that malignant transition had not occurred in this cancer. **5d)** Heatmap shows copy number variation at the single-cell level. The color scale for the copy number changes is shown at the right margin. The corresponding chromosomal locations are also shown. A total of 594 cells are represented. The clustering was performed as described in the Materials and Methods. Single-cell copy number variation analysis revealed two clusters, namely clusters A and NC (non-cancer). Cluster A features 1q + and 16q-structural variants and represents DCIS cells. Because cluster NC does not have a structural variant, it was considered to represent non-cancer cells from Case C.

To further examine the genomic clonality of this cancer, we performed single-cell copy number variation (scCNV) analysis. Cells in the cancer specimen were grouped into two clusters. Cluster A (542 cells), as shown in Fig. 5d, contained genomic aberrations at 1q+ and 16q−. These mutations, namely a gain of 1q and loss of 16q, are frequently observed in sporadic breast cancers^29,30^, especially histologically low-grade DCIS^31,32^, supporting the presumption that the lesion in Case C was low-grade DCIS. Contrarily, cluster NC (64 cells) had no genomic aberrations, and thus, they were considered non-cancer cells. Note that the proportions of cancer and non-cancer cells based on the morphological information (approximately 90 and 10%, respectively) were consistent with those observed via scCNV analysis (cluster A, 89%; cluster NC, 11%). Significantly, no subclone was detected in cluster A, indicating the absence of additional diversification of the cancer cells. These results differed from those for a malignant lesion (Case 7, which was also subjected to scCNV analysis) harboring *HER2* gene amplification and a microinvasion (Supplementary Fig. S8).

Together, the DCIS lesion of Case C did not harbor any genomic or transcriptomic alteration leading to the malignant transition, such as EMT and angiogenesis, which were observed in patients for *GATA3* mutation or downregulation, supporting the presumption that the lesion was possibly false DCIS. From the aforementioned clinical and molecular analyses, we propose the five most critical markers for the new classification of DCIS as follows: age less than 45 years, *HER2* amplification, *GATA3* mutation, or downregulation, PgR protein negativity (for high-risk and true DCIS), and *PIK3CA* mutation positivity (for false DCIS).

## Discussion

In this study, we identified the five most critical markers for classifying DCIS, namely age less than 45 years, *HER2* amplification, *GATA3* mutation positivity, *PIK3CA* mutation negativity, and PgR protein negativity, based on an analysis of the integrated clinical features of 431 patients with DCIS, whole-exome sequencing of a discovery cohort of 21 patients with DCIS, and targeted deep sequencing of a validation cohort of 72 patients with DCIS. We further analyzed the contribution of the extracted risk factors to the progression of invasive cancer *in vivo* using STseq and scDNA-seq. As the molecular bases, we found that aberrant *GATA3* function triggers the upregulation of EMT and angiogenic pathways at an early stage in the malignant transition of DCIS cells. PgR downregulation is likely a consequence of aberrant *GATA3* function accompanied by the upregulation of EMT-related pathways. *PIK3CA* mutation had a monoclonal structure, in which the expression modules were required specifically for proliferation pathways but not for EMT or invasive pathways, suggesting that *PIK3CA*-mutated DCIS is less likely to progress to IDC. There have been limited studies investigating *GATA3* mutation in DCIS^33^, and the prognostic consequence of the mutation remains to be clarified^34,35^. In the current study, *GATA3* mutation was identified as a key molecular marker for high-risk DCIS. STseq revealed that aberrant *GATA3* function, either through genomic mutations or transcriptomic downregulation, triggers the pivotal pathways of malignant transition that are expected to promote the progression of DCIS to IDC. *GATA3* promotes the differentiation of luminal cells and inhibits infiltration and metastasis of breast cancer cells^36–40^. Mutation of *GATA3*, which occurs at a much higher incidence in IDC^41^, is believed to abolish this critical function and promote the progression of cells to a more advanced stage. *GATA3* mutation may therefore play critical roles in the migration and invasion of cancer cells but not in the initial carcinogenesis. In clinical practice, examination of *GATA3* mutation is not always possible. In such cases, our results suggest that routine measurement of PgR protein expression would generally predict the functionality of GATA3 in patients with ER-positive DCIS.

Mutation of *PIK3CA* is crucial for breast carcinogenesis^42,43^. However, its mutation is correlated with a favorable prognosis in IDC^26^. It was also recently reported that mutation of the *PIK3CA* kinase domain and the absence of copy number gains in DCIS protect against progression to invasive cancer^44^. Our results that DCIS cells with *PIK3CA* mutations possess a monoclonal structure in which expression modules were required specifically for proliferation pathways support previous findings and indicate that *PIK3CA* mutation should be included as a marker for predicting a lower risk of progression to IDC and a reassessment of treatment options. A logical interpretation is that sequential mutation of *GATA3* facilitates the acquisition of invasive characteristics following *PIK3CA* mutation to promote tumor initiation. However, this is not likely for all lesions because there were several lesions with *GATA3* mutations but not *PIK3CA* mutations and vice versa. Whether *GATA3* and *PIK3CA* mutations are mutually exclusive initiating events at the single-cell level remains to be clarified.

We revealed that conventional clinicopathological factors are also critical for predicting the risk of IDC development, including age (≥45 years vs. <45 years), *HER2* status (negative vs. positive), and the combination of age and *HER2. HER2* amplification is a well-known poor prognostic factor in IDC in the absence of HER2-targeting therapy^45^. However, the frequency of *HER2* amplification differs between DCIS and IDC^46–50^, and whether *HER2* amplification is a risk factor for DCIS remains controversial^51^. Our results indicate that *HER2* positivity and age <45 years should be considered the major markers for high-risk DCIS.

In conclusion, we selected age <45 years, *HER2* amplification, *GATA3* mutation positivity, *PIK3CA* mutation negativity, and PgR protein negativity as the most reliable markers for predicting the risk of progression of DCIS to IDC, but we could not identify other substantial factors in our exhaustive genetic analyses. One limitation of this study was that samples from patients with no treatment history to confirm the outcome of DCIS are hardly available at present. However, clinical studies of DCIS observing its natural history in the absence of treatment, including ongoing trials^12–14^, may provide an accurate algorithm for discriminating high- and low-risk DCIS using markers including that proposed in this study to avoid unnecessary treatments.

## Methods

### Ethical approval

This study was approved (approval number: 2297-i103) by the Clinical Ethics Committee of St. Marianna University, and a waiver of consent was granted for the use of archival clinical samples from the Department of Pathology.

### Clinicopathological data

Clinicopathological data were obtained from 431 consecutive patients with DCIS who underwent surgery at University Hospital, St. Marianna University School of Medicine between 2007 and 2012. The detailed clinicopathological data are provided in SourceData1 and Supplementary Tables S1.

### Exome library construction and whole-exome sequencing

For the discovery cohort, we selected formalin-fixed, paraffin-embedded (FFPE) tissue samples from 21 patients with pure DCIS (primary tumor) and four patients with IDC (relapse tumors). In each case, epithelial areas in DCIS and normal epithelial tissue were identified via hematoxylin and eosin staining of the corresponding cryosections. Epithelial areas of interest were confirmed via histological assessment of each case by two histopathologists. Microdissected tissues (10 μm thick) using the Roche Automated Tissue Dissection System (Roche) were used to selectively isolate DCIS and non-tumor cells for whole-exome analysis, reducing the extent of contamination by stromal cells(Supplementary Fig. S2). DNA from the microdissected tissues was extracted using QIAamp DNA Mini and Micro kits (QIAGEN, Crawley, UK). DNA was tested for quality using a Tape Station (Agilent Technologies), and the concentration was assessed using a Bioanalyzer 2100 (Agilent Technologies).

Next-generation sequencing DNA libraries were prepared for whole-exome sequencing using 100–200 ng of DNA. In brief, whole-exome libraries were prepared using a SureSelect XT HS and XT Low Input Target Enrichment Kit (Agilent, UK) and OneSeq SS 300kb Backbone + Human All Exon V7 capture library (Agilent Technology) following the manufacturer’s guidelines. Whole-exome libraries were sequenced using 100-bp paired-end runs on the Illumina HiSeq 2500/3000 system (Illumina).

The sequencing data were analyzed using a custom pipeline. In brief, sequencing reads were aligned to the human genome (hg 19) using the Burrows-Wheeler Aligner Mem (ver 0.7.17)^52^. Duplications were marked using Picard Tools V2.18.25 (http://broadinstitute.github.io/picard). Insertion–deletion realignment and base recalibration were achieved using GATK v.4.0.12-0^53^.

The somatic variants were detected using an ensemble approach with two variant callers: MuTect2^54^ and Genomon pipeline^55^. The variant annotation was performed using ANNOVAR^56^. To obtain the final set of mutation calls, we used a two-step approach: 1) to reduce false-positive calls, the mutant variant frequency must be at least 4% of total reads; 2) to remove any spurious variant calls arising as a consequence of sequencing artifacts, we checked bam reads using Integrative Genomics Viewer (http://www.broadinstitute.org/igv/). Only variants with the following functional classification were considered in this study: non-synonymous single nucleotide variants, stop gain mutation, and frameshift mutation. The full list of non-synonymous variants is provided in SourceData6.

### Target sequencing library construction and deep targeted sequencing

For the validation study, we conducted deep targeted sequencing. We selected 72 patients with pure DCIS and available FFPE tissue. Because of the limited number of specimens, only tumor samples were analyzed. To enrich DCIS cells, manually macrodissected tissues were used. DNA from the macrodissected tissues was extracted using QIAmp DNA Mini and Micro kits and tested for quality using the Tape Station (Agilent Technologies), and the concentration was assessed using a Bioanalyzer 2100.

Next-generation sequencing DNA libraries were prepared for target sequencing using 100–200 ng of DNA. In brief, target sequence libraries were prepared using a SureSelect XT HS and XT Low Input Target Enrichment Kit and custom sequencing panel following the manufacturer’s guidelines. A custom sequencing panel containing 180 genes was designed (full list in SourceData3). This panel targeted genes that were duplicated in the whole-exome sequencing study of the preceding 21 patients including *GATA3, PIK3CA*, and breast cancer related genes. The total region size of the custom panel was 1.5 Mbp. Target sequence libraries were sequenced using 100-bp paired-end runs on the Illumina Novaseq. The sequencing data were analyzed using a custom pipeline as described in the “Exome library construction and whole-exome sequencing” subsection. The full list of non-synonymous variants is provided in SourceData7.

### Subclonal analysis

The number of subclones contributing to each sample was estimated using the PyClone program version 0.13^23^. We used the sequenza package to define the CNV variation from whole-exome sequencing data^57^. We set the major and minor allele copy numbers as those obtained from the sequenza program, allowing clustering to simply group clonal and subclonal mutations. PyClone was operated with 10,000 iterations and default parameters. PyClone results were analyzed using CloneEvol to infer and visualize clonal evolution as described previously^23^.

### STseq using Visium

We selected three cases for STseq using Visium. The clinical information of each patient is provided in Supplementary Table S3.

Each sample was collected immediately after surgical removal and embedded in optimal cutting temperature (OCT) compound (TissueTek Sakura) in a 10 mm × 10 mm cryomold at −80°C until use. Frozen samples embedded in OCT compound were sectioned at a thickness of 10 μm (Leica CM3050 S). Libraries for Visium were prepared according to the Visium Spatial Gene Expression User Guide (CG000239_VisiumSpatialGeneExpression_UserGuide_Rev_A.pdf). Tissue was permeabilized for 6 min, which was identified as the optimal time in tissue optimization time course experiments.

Libraries were sequenced on a NovaSeq 6000 System (Illumina) using a NovaSeq S4 Reagent Kit (200 cycles, 20027466, Illumina) at a sufficient sequencing depth (approximately 700 million to 1.2 billion reads per sample).

Sequencing was performed using the following read protocol: read 1, 28 cycles; i7 index read, 10 cycles; i5 index read, 10 cycles; read 2, 91 cycles. Raw FASTQ files and histology images were processed using Space Ranger software v1.0.0 (https://support.10xgenomics.com/spatial-gene-expression/software/pipelines/latest/installation). To visualize spatial expression using histological images, the raw Visium files for each sample were read into Loupe Browser software v4.0.0 (https://support.10xgenomics.com/spatial-gene-expression/software/downloads/latest).

To assign individual spots to tumor cells or cells that comprise the microenvironment, we compared the clusters morphologically annotated by a pathologist to data-driven spatial clusters using k-means clustering results provided by 10x Genomics Space Ranger software (Fig. 3a, Fig. 5a, Supplementary Fig. S7).

We extracted reads with confirmed *GATA3* mutation in whole-exome sequencing from the Visium alignment results, and we identified spots with *GATA3* mutations (Fig. 3b, Supplementary Fig. S6).

### Differential expression analysis and pathway analysis of each Visium case

We performed differential expression analysis and pathway analysis at the spot level using our Visium data.

In Case A, we compared 46 spots with *GATA3* mutations (red color in Fig. 3b) to 376 spots without *GATA3* mutations (green color in Fig. 3b). Count data in raw Visium files were used. DEGs were identified using the TCC package^27^ and a filtering threshold of FDR < 0.01. The TCC package was also used to generate a MA plot to visualize DEGs. Metascape^28^ (http://metascape.org) was used to perform pathway enrichment analysis. The top 100 genes according to FDR were subjected to Metascape, which was performed on two groups of gene sets, namely the MSigDB v5.1 Hallmarks gene set collection (HALLMARK)^58^ and Kyoto Encyclopedia of Genes and Genomes (KEGG). Pathways were considered statistically significant at P ≤ 0.05. To assess whether differences in mRNA expression between the spots with and without *GATA3* mutations using an unpaired *t*-test (Fig. 3e), row count data were normalized as counts per 10,000 by dividing each spatial spot column by the sum of its counts and multiplying by 10,000.

In Case B, to focus on the gene expression changes in DCIS cells, we manually selected 188 spots that were morphologically located in the region of the milk ducts. Unsupervised hierarchical clustering based on Spearman’s distance and Ward’s linkage was used to build a tree relating the clusters. Hierarchical clustering analysis of these spots identified three apparent clusters (clusters 1–3). The 48 spots of cluster 1 overlapped with the location of DCIS cells that were about to invade on the basis of their morphology. Meanwhile, the 101 spots of cluster 2 were located in the proximal ducts that were anatomically the same region as the spots of cluster 1. We compared the 48 spots in cluster 1 (red color in Fig. 4b) to the 101 spots in cluster 2 (green color in Fig. 4b). Count data in raw Visium files were used. DEGs were identified using the TCC package and a filtering threshold of FDR < 0.01. The TCC package was also used to generate a MA plot to visualize DEGs. Metascape was used to perform pathway enrichment analysis using the HALLMARK and KEGG gene sets. Pathways were considered statistically significant at P ≤ 0.05. To assess differences in mRNA expression between the spots in clusters 1 and 2 using an unpaired *t*-test (Fig. 4e), row count data were normalized as counts per 10,000 by dividing each spatial spot column by the sum of its counts and multiplying by 10,000.

### Co-expression network analysis of Visium Case C

Case C exhibited a monotonous gene expression pattern, which was also consistent with the morphologically monoclonal structure of this cancer (Fig. 5a). The spots were roughly separated into two clusters via un-hierarchical k-means clustering. They almost completely overlapped with morphological cancer (347 spots) and non-cancer cells (151 spots). Thus, we compared the 347 spots with cancer cells (blue color in Fig. 5a) with the 151 spots with stroma cells (orange color in Fig. 5b). DEG analysis using the TCC package (as described in the differential expression analysis and pathway analysis of each Visium case subsection) between the cancer and non-cancer spots revealed 2364 DEGs (data not shown).

To group related genes into gene modules (clusters) based on their co-expression patterns, we used WGCNA^59^. For WGCNA, 508 DEGs that were upregulated in cancer spots were used to construct a co-expression network. Using the pickSoftThreshold function with a fit value exceeding RsquaredCut, which was 0.8, the power (β) parameter was inferred to be 3. WGCNA could not confidently assign 439 genes to any of the modules because they displayed little correlation with any other gene. These uncorrelated genes were designated as module 0 and excluded from the rest of the analysis. We conducted subnetwork analysis to identify the hub genes in each module. Pathway enrichment analysis of these hub genes using Metascape revealed that genes involved in the estrogen response and p53 pathways were enriched in module 1.

### scDNA library preparation and scDNA-seq

Tissue biopsies were obtained from surgically resected primary DCIS samples. Samples were washed in PBS (Wako 045-29795), mechanically dissociated using a razor blade, and digested in DMEM (Wako 041-29775) containing collagenase Type P, 2 mg/mL (Roche 11213857001). Cellular debris and aggregates were filtered using a 40-μm cell strainer (CosmoBio) prior to scDNA-seq. Single-cell suspensions were loaded into the Chromium 10x device according to the standard protocol provided with the Chromium Single-Cell DNA Reagent Kits (10x Genomics).

For droplet-enabled scDNA-seq, we used the 100-bp paired-end Illumina HiSeq3000. Sequencing data was processed using cellranger-dna-1.1.0 (refdata-GRCh38-1.0.0.), which automated sample demultiplexing, read alignment by bwa mem with -M options, correction of GC bias by fitting to the quadratic function that minimizes the entropy of the read count histogram per 20 kbp, CNV calling with a 20-kb bin size, and report generation. We excluded noisy cells and cells with ploidy <1.9 or >2.0 (Her2 >8 is an exception) from the cellranger-dna result.

A CNV heatmap was plotted using the pheatmap function in R package, and using the Manhattan distance, we computed a hierarchical clustering using Ward’s minimum variance method. To identify subclusters from CNA Data, the optimal ‘k’ (number of clusters) was determined using the elbow Method and Silhouette Method in R package ‘factoextra,’ viz_nbclust function (FUN = hcut, k.max =15). The tree was cut into k clusters to determine the number of clones and optimized manually.

### Immunohistochemistry and measurement of protein expression

Paraffin tissue sections measuring 4 μm on coated slides were deparaffinized using routine techniques. The expression of ER, PR, and HER2 was determined using standard immunohistochemical and fluorescent in situ hybridization techniques for clinical analyses.

HER2 overexpression was analyzed according to the American Society of Clinical Oncology and College of American Pathologists (ASCO-CAP) 2013 recommendations, specifically, HER2 staining was considered positive if the circumferential membrane staining was complete, intense, and present in more than 10% of tumor cells (HER2 score 3+) or if circumferential membrane staining was incomplete and/or weak/moderate and present in more than 10% of tumor cells (HER2 score 2+) if HER2 overexpression could be confirmed via fluorescence in situ hybridization. HER2 staining was considered negative when incomplete membrane staining was faint/barely visible in >10% of the tumor cells (HER2 score 1+) or when no staining was observed (HER2 score 0)^60^.

### Statistical analyses

Statistical analyses were performed using GraphPad Prism v8.0 and R version 3.5.0. Fisher’s exact was employed for comparisons of unordered categorical variables. Student’s *t*-test was utilized to compare continuous variables and ordered categorical variables.

Relapse-free survival was defined as the time from the date of surgery to that of recurrence or the last contact. Survival curves were constructed using the Kaplan– Meier method and compared using the log-rank test. Cox proportional hazard regression models, including unadjusted models and models adjusted for available prognostic clinical and genomic covariates, were constructed to calculate HRs and 95% CIs.

Statistical significance was accepted at P < 0.05 (two-sided, unless otherwise indicated).

## Data Availability

All sequencing data and pathological images for STseq have been deposited in the DNA Data Bank of Japan under the accession number JGAS00000000202.

## Acknowledgements

We thank all patients who participated in this study.

We thank K. Imamura, K. Abe, Y. Ishikawa, M. Konbu, E. Kobayashi, E. Ishikawa, and T. Horiuchi for their technical assistance.

The authors would like to thank Enago (www.enago.jp) for the English language review.

## Authors’ contributions

S.N. and Y.S. designed the study. Y. Kojima., A.M., and T.I. provided specimens.

I.M. and S.N. conducted the pathological review. S.N. performed the microdissection experiments. S.N. and Y. Kuze performed mutation calling and analyzed spatial transcriptome data, scCNV data, and clonal dynamics. Y. Kojima., A.M., T. Onishi, T.I., T.Y., K.T., A.S., M.S., and K.T. offered advice and reviewed the manuscript. S.N., T. Ohta, and Y.S. prepared the manuscript.

## Supplementary Information

**Supplementary Fig. S1.**
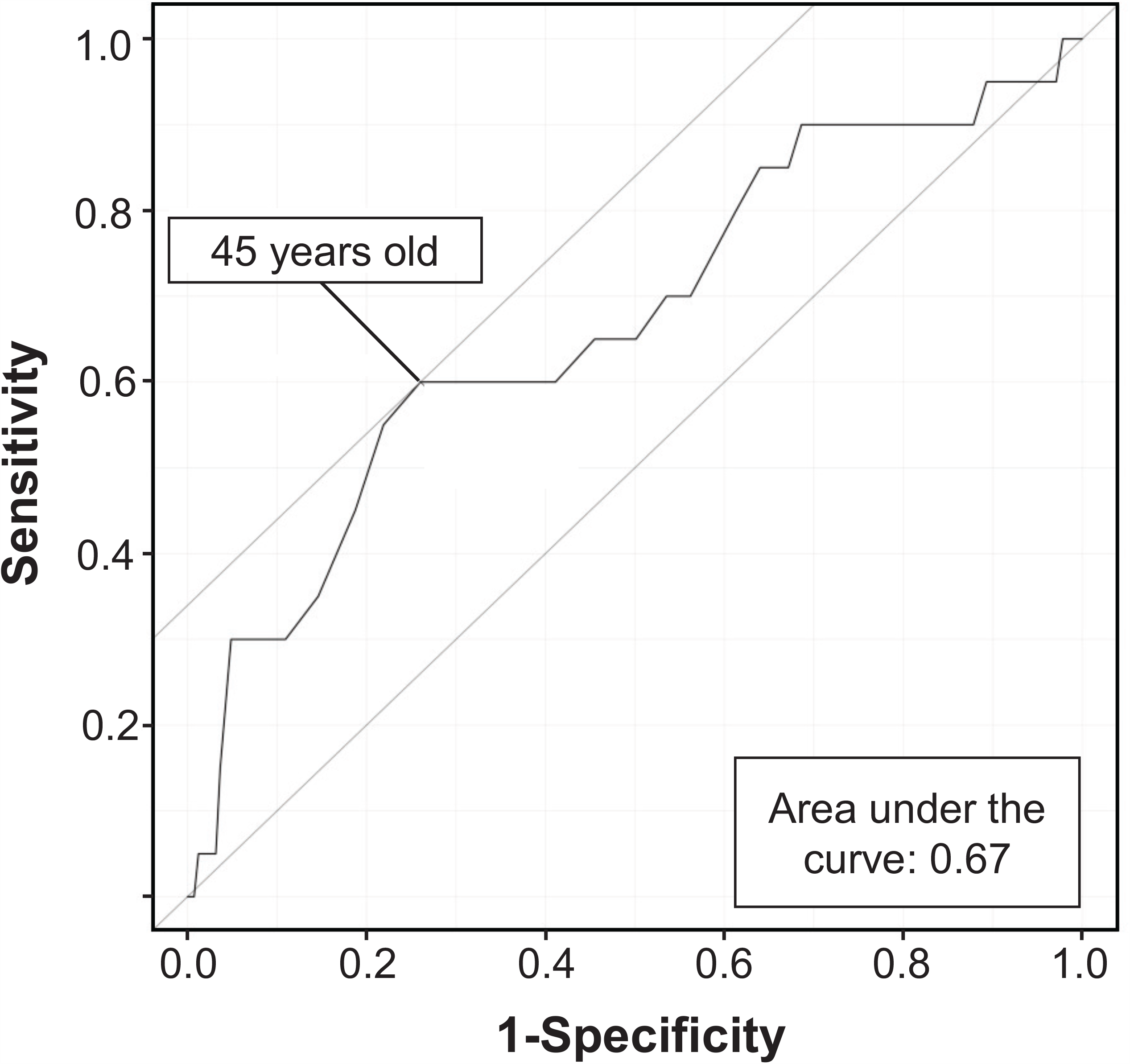
The receiver operating characteristic (ROC) curve for age. The resulting ROC curve, with an area under the curve (AUC) of 0.67, reveals a poor prognosis. According to the AUC, we selected 45 years as the cutoff (<45 years vs. ≥45 years).

**Supplementary Fig. S2.**
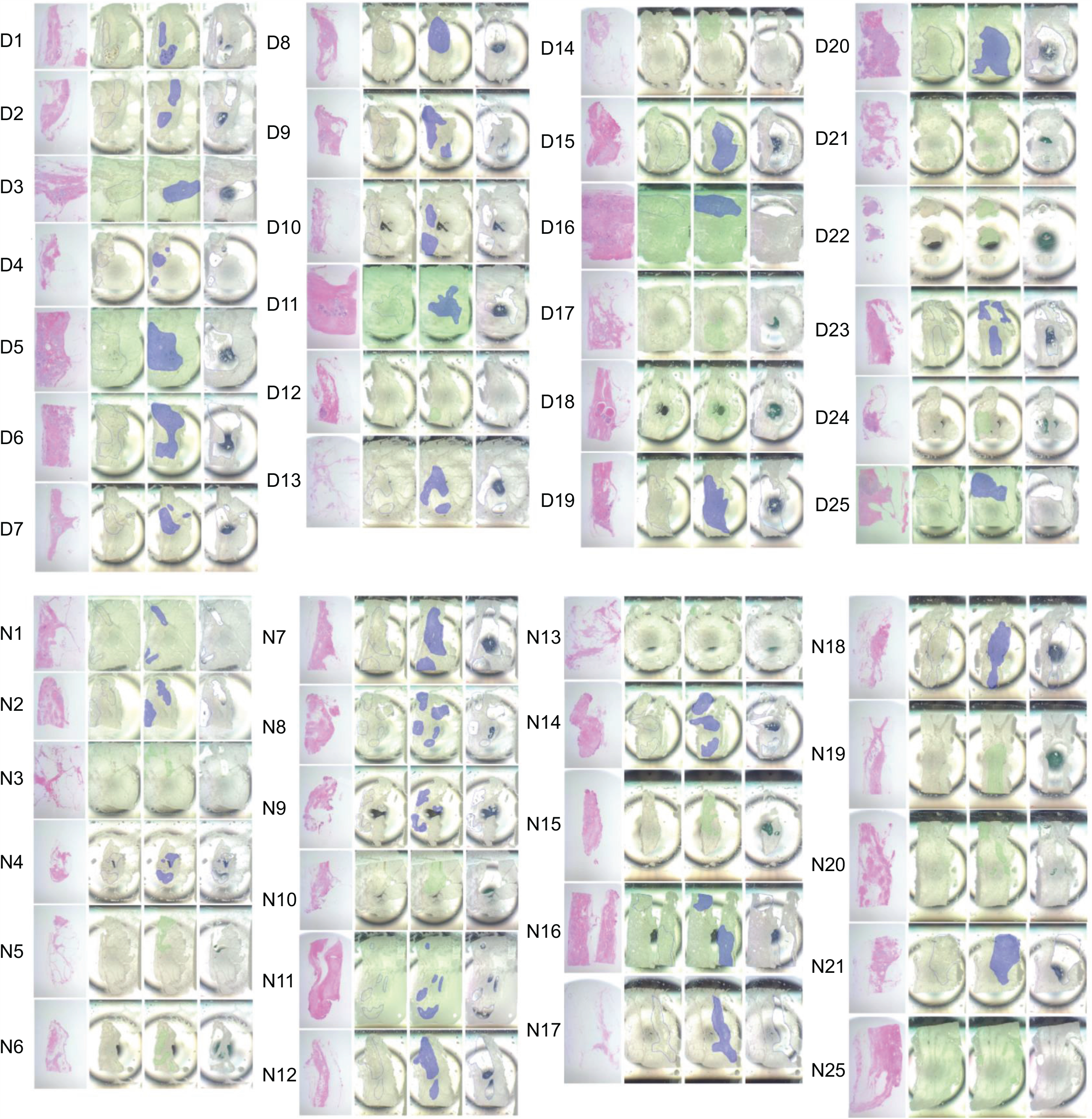
Microdissection of 25 pure ductal carcinoma in situ (DCIS) specimens in the discovery cohort. Specimens were subjected to careful microdissection to avoid contamination with cells. The first panel from the left shows hematoxylin and eosin staining. The second panel shows the specimens before microdissection. The third panel shows the target region of microdissection. The rightmost panel shows the remaining specimen after microdissection. D, DCIS; N, normal

**Supplementary Fig. S3.**
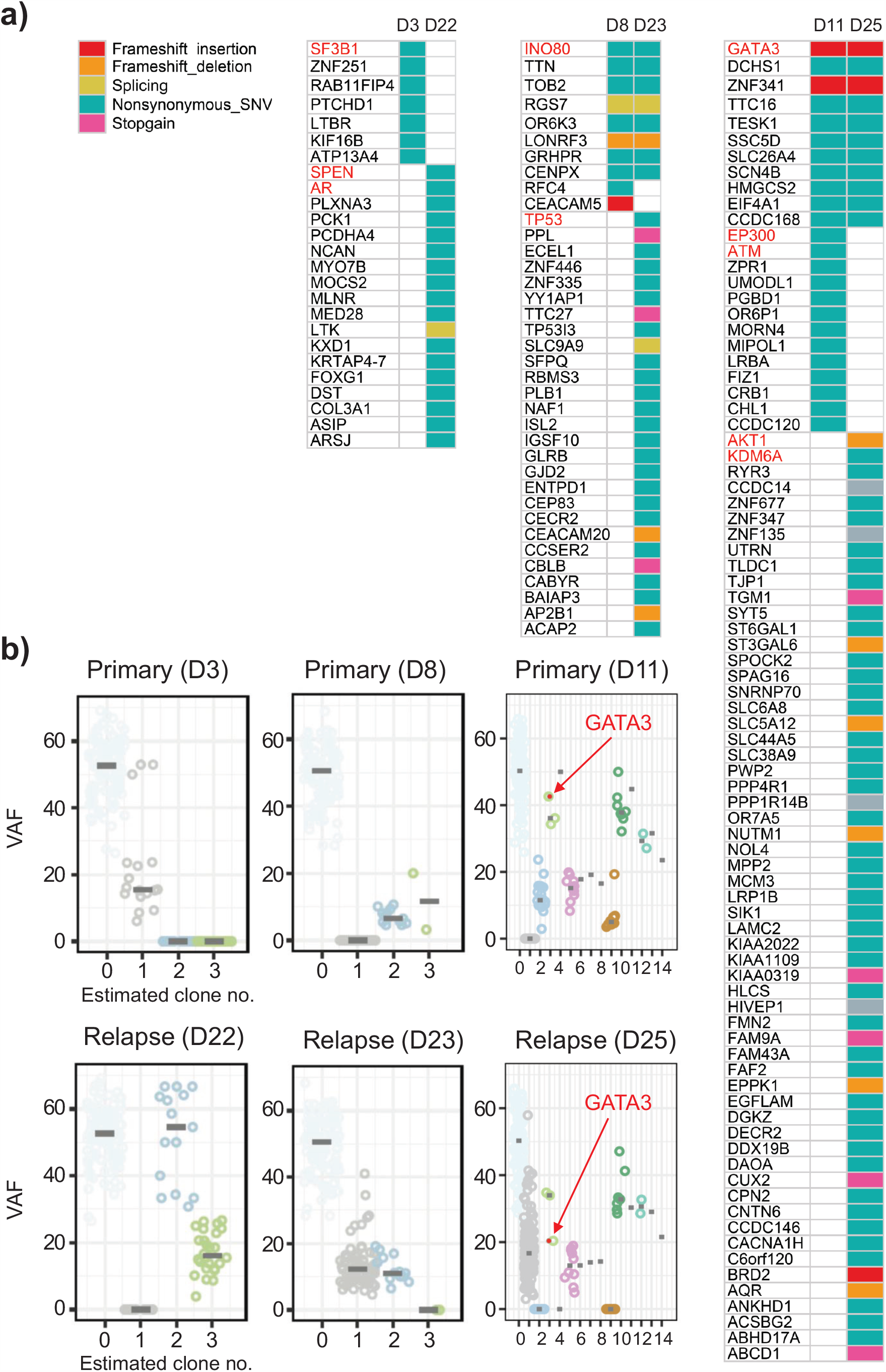
Comparison of mutations between primary and relapse lesions. **S3A:** Comparisons of mutations via whole-exome sequencing of primary (D3, D8, and D11) and matched relapse lesions (D22, D23, and D25) are shown. Well-known genes that cause breast cancer are colored red. No foundation mutation was shared by Cases D3 (primary lesion) and D22 (relapse lesion). Therefore, we assumed that recurrence in D3 was indeed a *de novo* breast cancer (new primary) rather than a recurrence of the primary ductal carcinoma in situ lesion. Contrarily, cases D8 and D11 (matched relapse lesions were labeled D23 and D25, respectively) shared foundation mutations. This suggested that a subclone in the primary lesion also existed in the relapse lesion. Therefore, we assumed that the recurrences in D8 and D11 were indeed relapses of breast cancer (true recurrence). **S3B:** The results of subclone analysis performed using PyClone software are shown.

**Supplementary Fig. S4.**
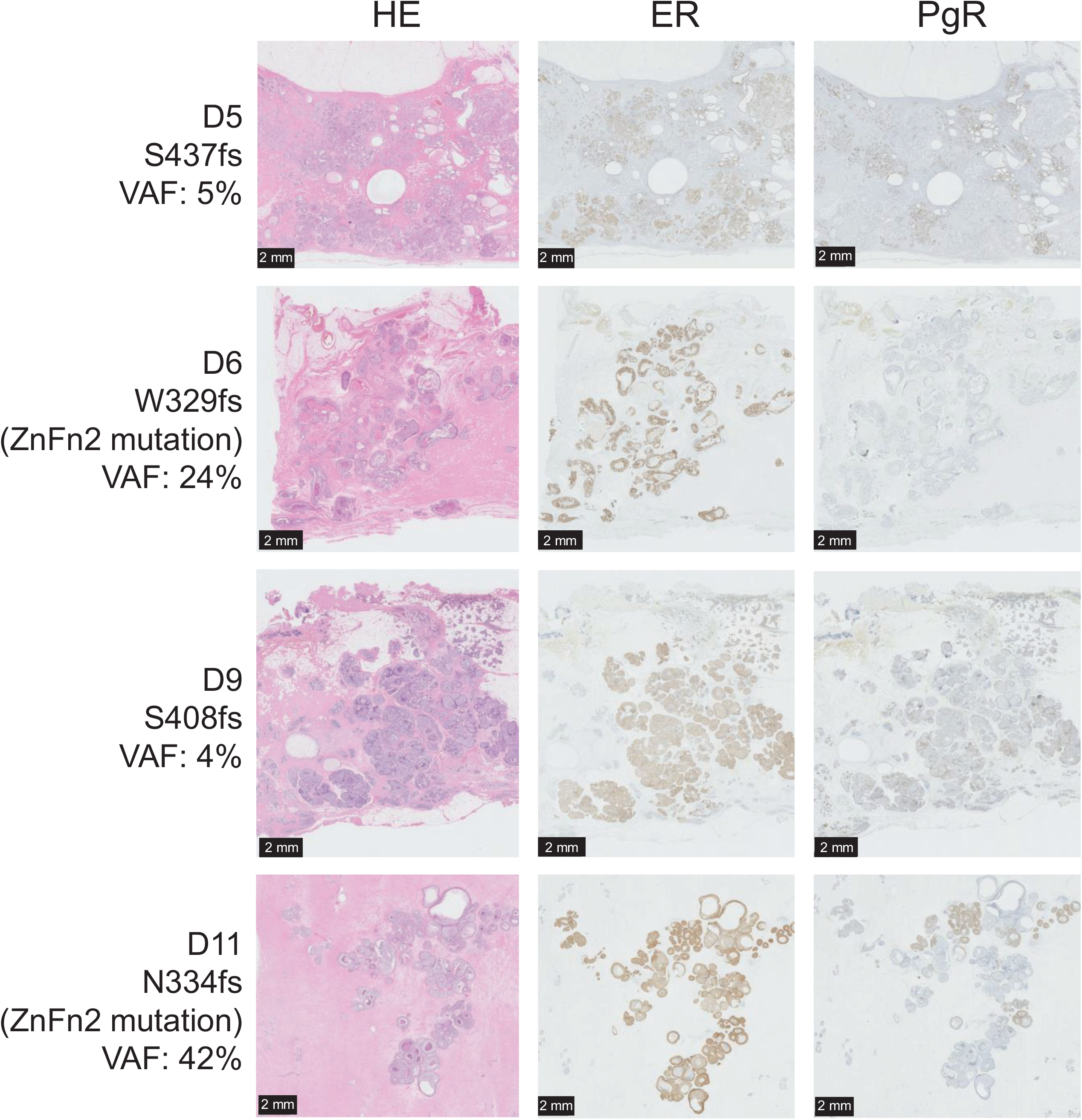
Progesterone receptor (PgR) expression in *GATA3* mutation-positive ductal carcinoma in situ. The panel presents hematoxylin and eosin, estrogen receptor (ER), and PgR staining for patients D5, D6, D9, and D11 with *GATA3* mutation. Although ER expression was positive, PgR expression was downregulated in all cases. Reflecting the variant allele frequency of *GATA3* mutation, some heterogeneity in PgR expression was observed. Scale bars: 2 mm.

**Supplementary Fig. S5.**
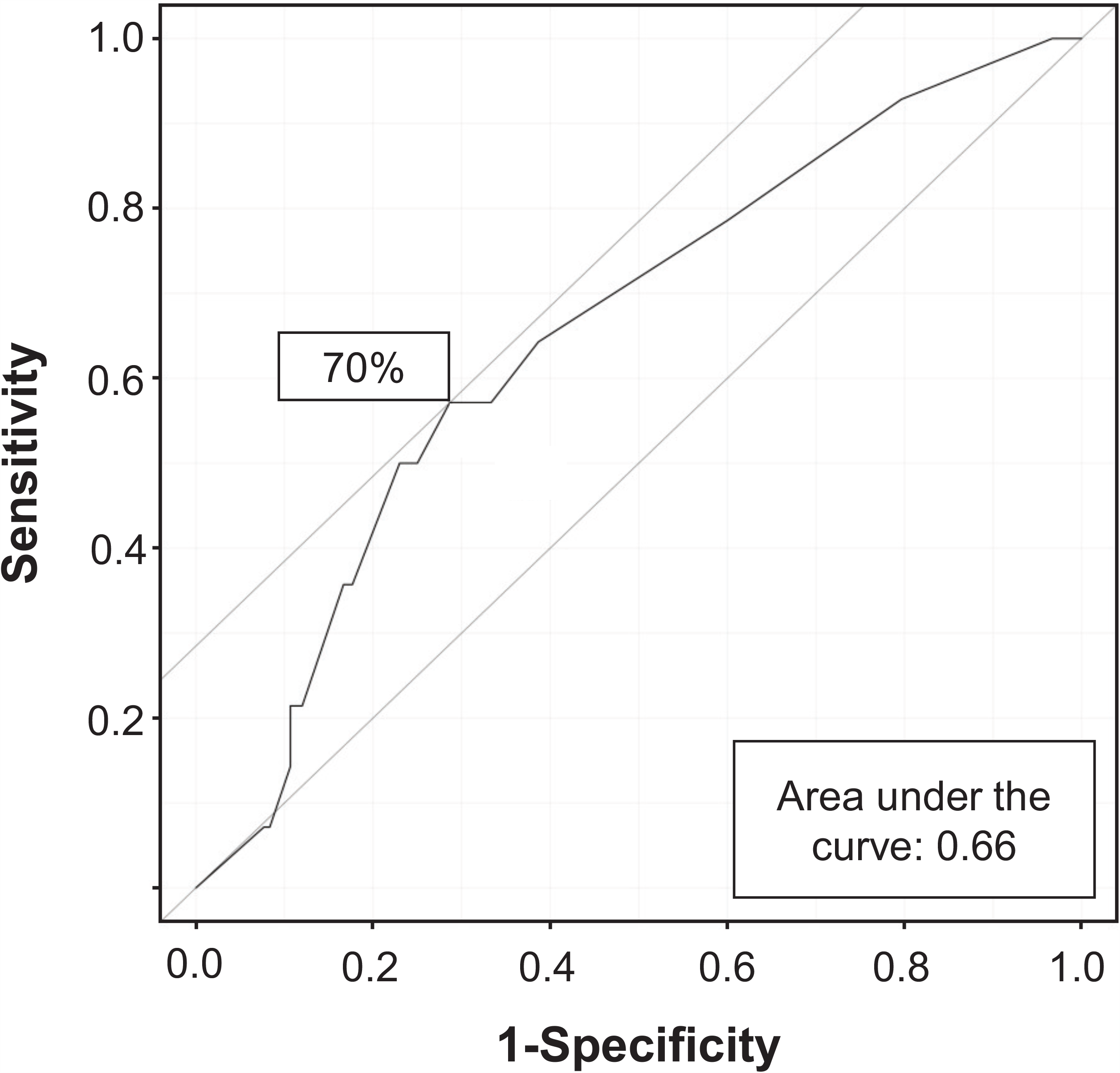
Receiver operating characteristic (ROC) curve for progesterone receptor (PgR) expression. The resulting ROC curve, which had an area under the curve of 0.66, denoted poor prognosis. Therefore, we decided that PgR expression in 70% or more of cells represented high expression, whereas expression in 60% or fewer cells indicated low expression.

**Supplementary Fig. S6.**
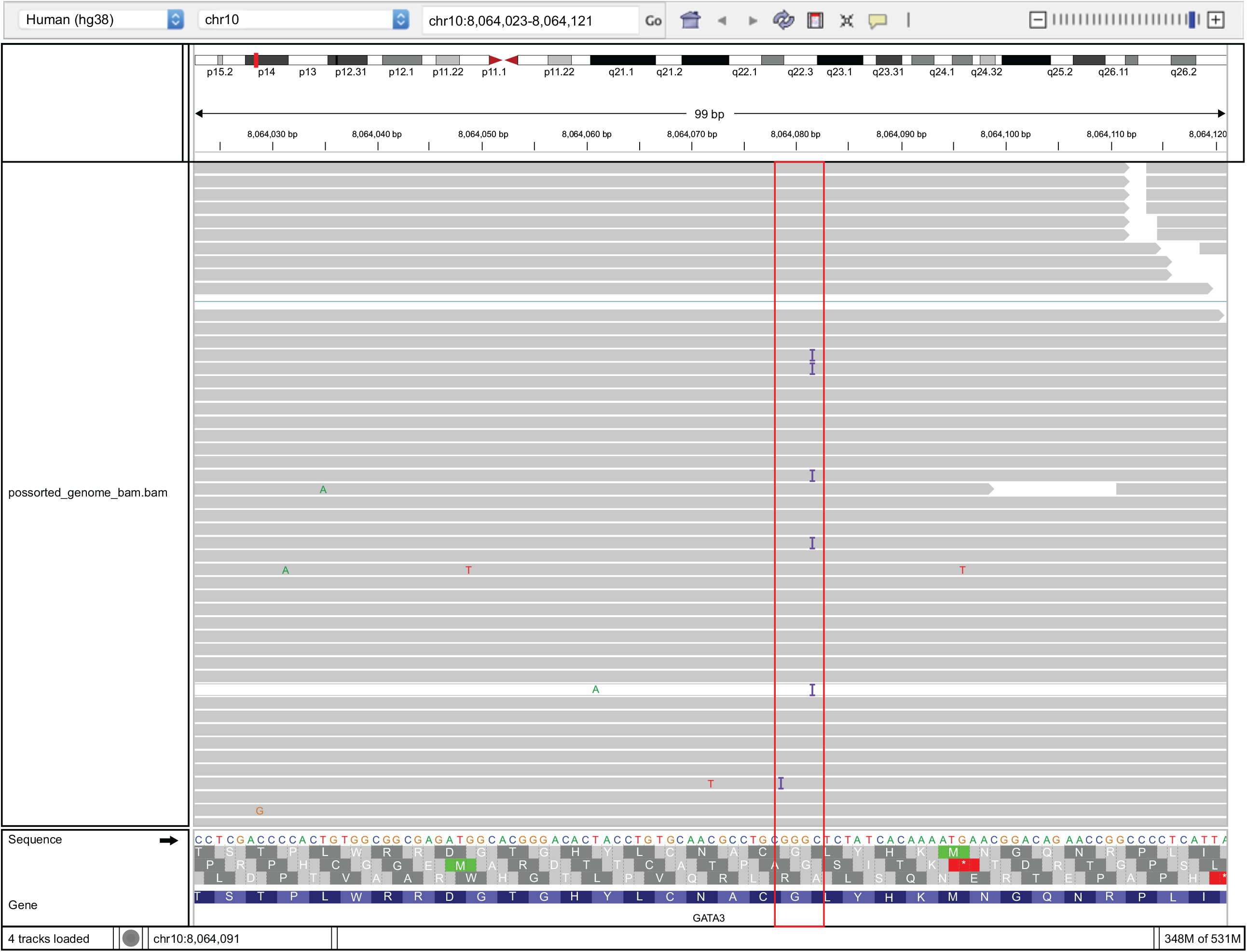
Selection of reads with *GATA3* mutation using the Integrative Genomics Viewer. Spatial transcriptome sequencing (STseq) reads on *GATA3* are shown. In Case A, the STseq reads revealed a *GATA3* mutation (exon4:c.865dupG:p.C288fs, indicated by a red square), and the mutation was located relatively near the 3′-end of mRNA.

**Supplementary Fig. S7.**
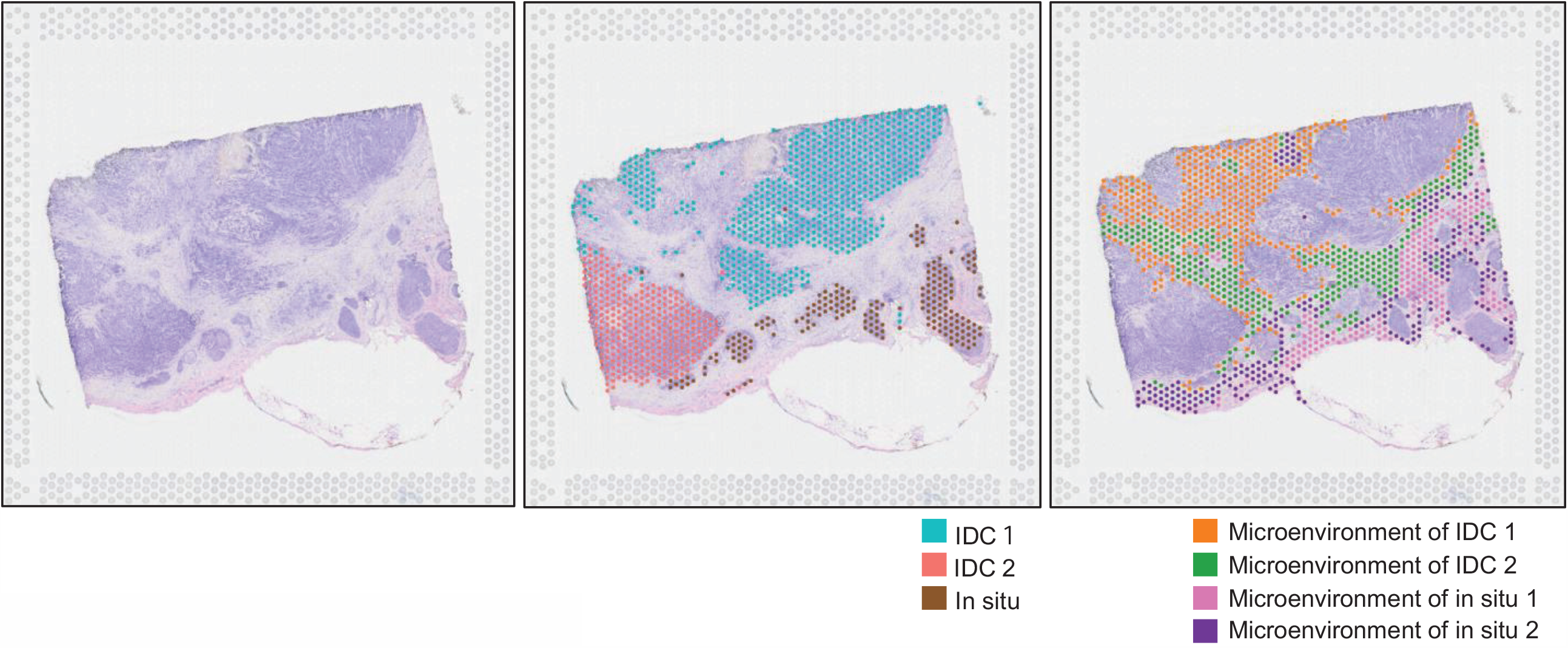
Visualization of the Visium results of Case B. Hematoxylin and eosin staining (left). Cancer spots were classified into three clusters via un-hierarchical k-means clustering (k = 9, middle). Non-cancer spots (microenvironment) were classified into four clusters (right).

**Supplementary Fig. S8.**
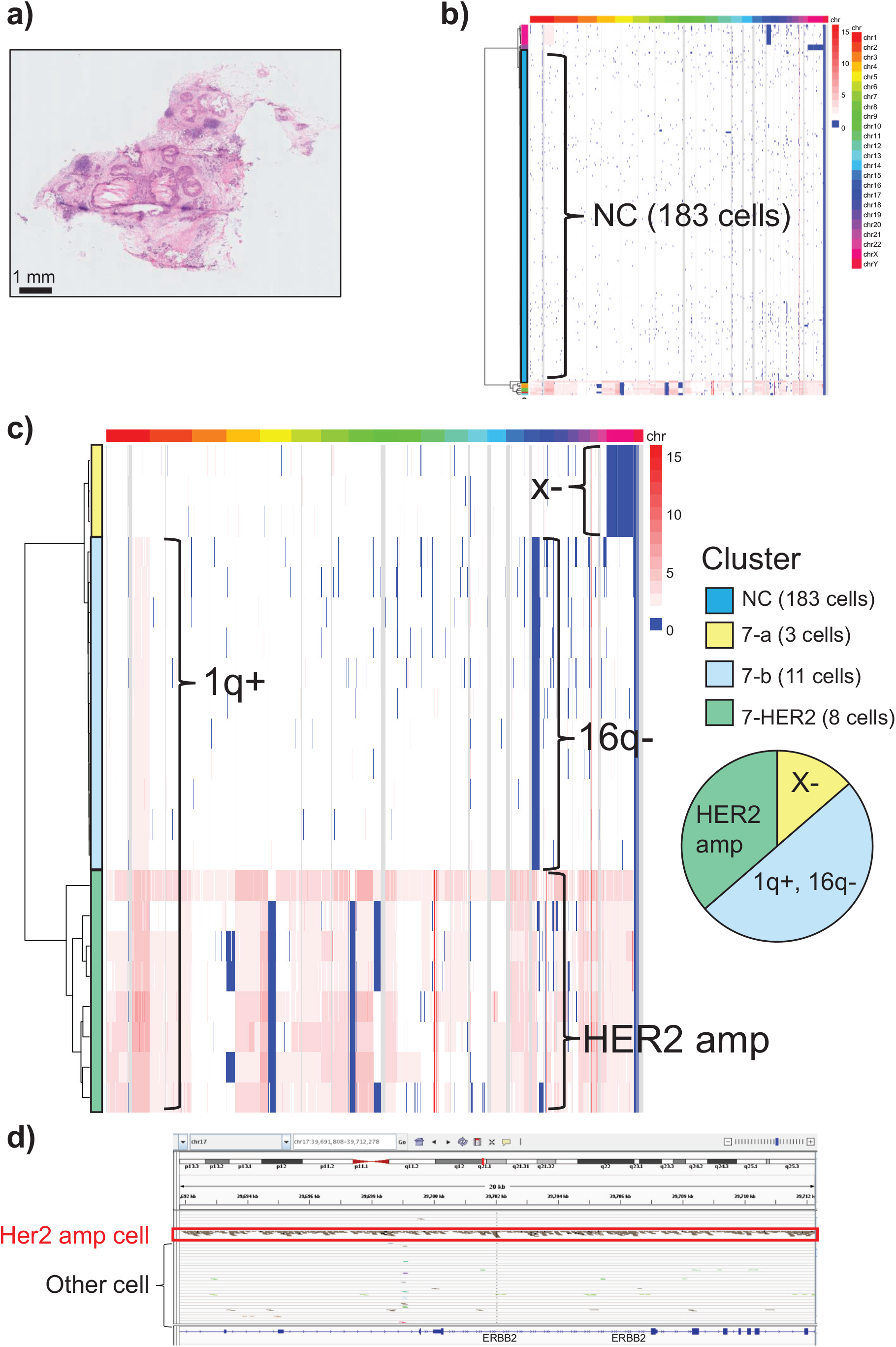
Single-cell copy number variation analysis of a malignant case (Case 7) **a)** Hematoxylin and eosin staining for Case 7. Case 7 was a 48-year-old woman (estrogen receptor, 0%; progesterone receptor, 0%; human epidermal growth factor receptor 2 immunohistochemistry score, 3+; Ki-67 index, 50%) who harbored a microinvasion. Therefore, this case was considered a typical case of malignancy. No mutation was detected in *PIK3CA* or *GATA3*. The scale bar is 1 mm. **b)** Heatmap presents the copy number variation at the single-cell level. The color scale for copy number changes is shown on the right margin. The corresponding chromosomal locations are also shown. A total of 205 cells are represented. The clustering was performed as described in the Materials and Methods. The generated clusters were designated as indicated. The non-cancer cell cluster was designated NC. The remaining cancer cells were further divided into subclusters as shown in (c). c) Magnification of the cancer populations in (b). The cancer cells were further divided into three major subclusters. The observed three major subclusters were designated clusters 7-a, 7-b, and 7-HER2. Subcluster 7-a (three cells) featured chrX−. Subcluster 7-b (11 cells) harbored 1q+ and 16q−. Subcluster 7-HER2 was further divided into subclones. Note the only subcluster 7-HER2 displayed genomic human epidermal growth factor receptor 2 (*HER2*) amplification. The pie chart on the right margin represents the composition of the cellular populations harboring the indicated genomic mutations. d) Single-cell DNA sequencing reads of the *HER2* region are shown using the Integrative Genomics Viewer. Red squares represent cells with *HER2* amplification.

**Supplementary Fig. S9.**
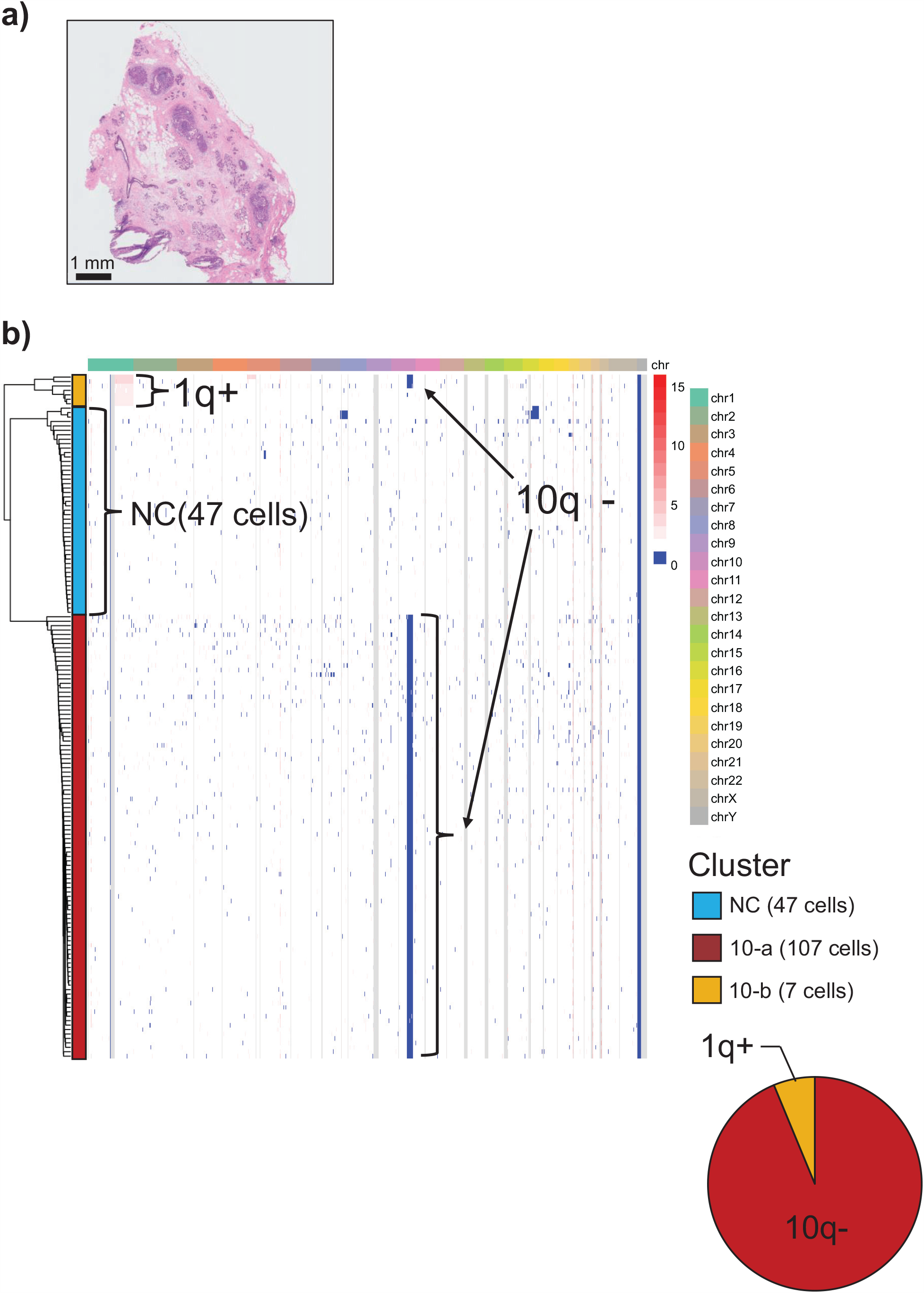
Single-cell copy number variation analysis in a patient without relapse (Case 10) **a)** Hematoxylin and eosin staining in Case 10. Case 10 was a 47-year-old woman (ER, 99%; PgR, 99%; human epidermal growth factor receptor 2 immunohistochemistry score, 2+ (FISH 1.1); Ki-67 index, 1-9%) who did not harbor a microinvasion. No mutation was detected in *PIK3CA* and *GATA3*. The scale bar is 1 mm. **b)** Heatmap presents the copy number variation at the single-cell level. A total of 161 cells are represented. The non-cancer cell cluster was denoted by NC. The cancer cells were further divided into two subclusters, including one major (subcluster 10-a) and one minor subcluster (subcluster 10-b). Subcluster 10-a (107 cells) harbored 10q−. Subcluster 10-b (seven cells) harbored 1q+. Because some clones in subcluster 10-b featured same structural variants as subcluster 10-a, all of the cancer cells were considered to comprise a single cluster. The pie chart on the right margin represents the composition of the cellular populations having the indicated genomic mutations.

**Supplementary Table S1.** Clinicopathologic characteristics of patients in the discovery and validation cohorts

| Characteristic | Discovery cohort<br>(N = 21) | Validation cohort<br>(N = 72) | Difference<br>(P) |
| --- | --- | --- | --- |
| Age at diagnosis |  |  |  |
| Median age (range), years | 42 (29–76) | 49 (34–76) | 0.3292 |
| ≥45 years, n (%) | 9 (42.9) | 47 (65.3) |  |
| <45 years n (%) | 12 (57.1) | 25 (34.7) |  |
| Management, n (%) |  |  |  |
| BCS | 10 (47.6) | 49 (68.1) | 0.2099 |
| Mastectomy | 4 (19.0) | 10 (13.9) |  |
| NSM | 7 (33.3) | 13 (18.0) |  |
| ER status, n (%) |  |  |  |
| Negative | 5 (23.8) | 20 (27.8) | 0.7875 |
| Positive | 16 (76.2) | 52 (72.2) |  |
| PgR status, n (%) |  |  |  |
| Negative | 6 (28.6) | 23 (31.9) | >0.9999 |
| Positive | 15 (71.4) | 49 (68.1) |  |
| HER2 amplification, n (%) |  |  |  |
| No | 13 (61.9) | 40 (55.6) | 0.8027 |
| Yes | 8 (38.1) | 32 (44.4) |  |
| Nuclear atypia, n (%) |  |  |  |
| Low grade | 6 (28.6) | 14 (19.4) | 0.0956 |
| Intermediate grade | 7 (33.3) | 43 (59.7) |  |
| High grade | 8 (38.1) | 15 (20.8) |  |
| Comedo necrosis, n (%) |  |  |  |
| No | 8 (38.1) | 30 (41.7) | 0.8066 |
| Yes | 13 (61.9) | 42 (58.3) |  |
| Median tumor size(range), cm | 3.0 (1.2–13.4) | 2.1 (0.1–10.5) | 0.04928 |

BCS, breast-conserving surgery; NSM, nipple-sparing mastectomy; ER, estrogen receptor; PgR, progesterone receptor; HER2, human epidermal growth factor receptor 2

**Supplementary Table 2.**

|  | PgR low | PgR high |  |
| --- | --- | --- | --- |
| GATA3 mt (+) | 4 | 0 | 4 |
| GATA3 mt (−) | 3 | 9 | 12 |
|  | 7 | 9 | 16 |

mt, mutation; PgR, progesterone receptor

**Supplementary Table S3.**
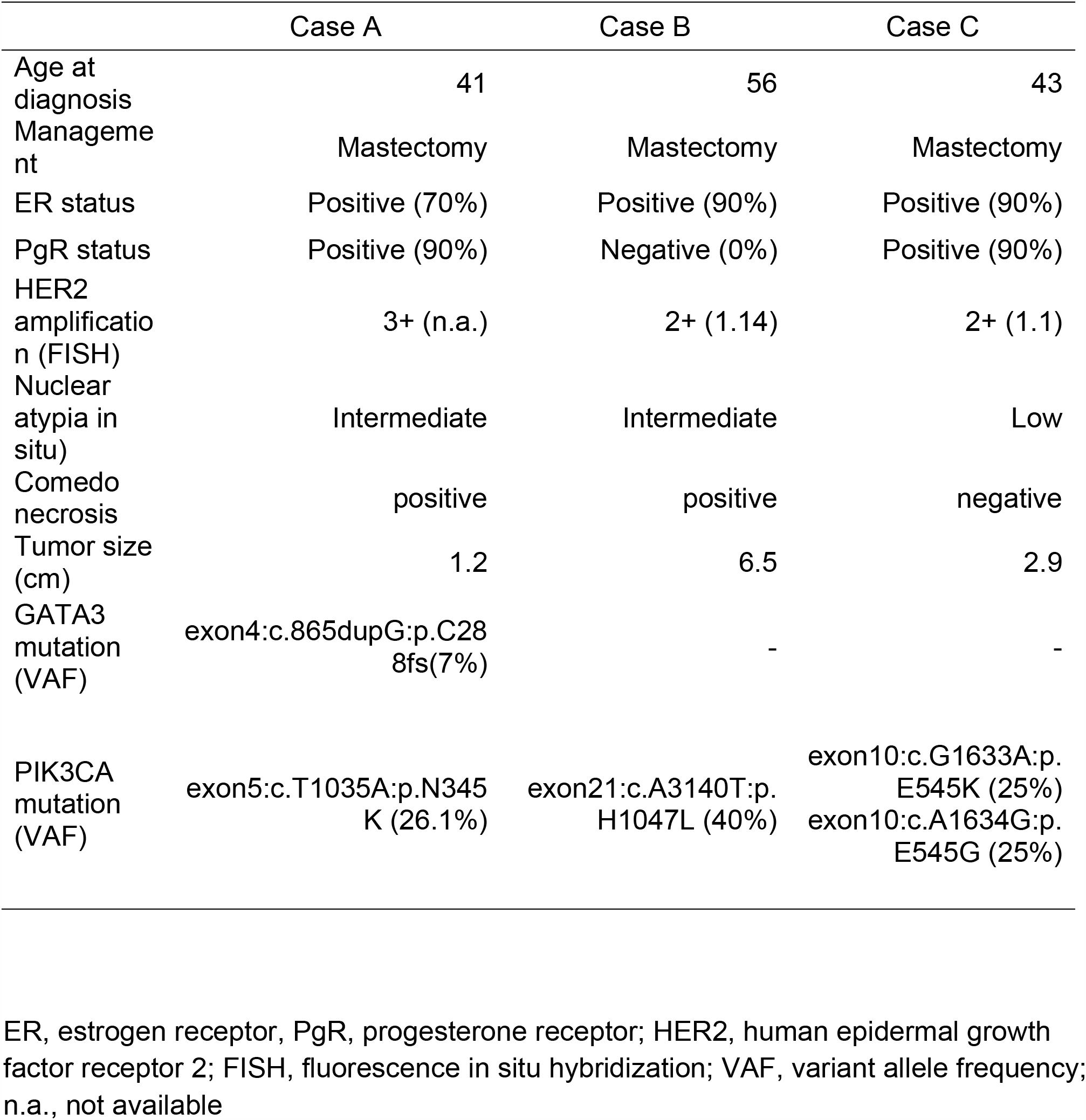
Clinical information of Visium cases

|  | Case A | Case B | Case C |
| --- | --- | --- | --- |
| Age at diagnosis | 41 | 56 | 43 |
| Management | Mastectomy | Mastectomy | Mastectomy |
| ER status | Positive (70%) | Positive (90%) | Positive (90%) |
| PgR status | Positive (90%) | Negative (0%) | Positive (90%) |
| HER2 amplification (FISH) | 3+ (n.a.) | 2+ (1.14) | 2+ (1.1) |
| Nuclear atypia in situ) | Intermediate | Intermediate | Low |
| Comedo necrosis | positive | positive | negative |
| Tumor size (cm) | 1.2 | 6.5 | 2.9 |
| GATA3 mutation (VAF) | exon4:c.865dupG;p.C288fs(7%) | - | - |
| PIK3CA mutation (VAF) | exon5:c.T1035A:p.N345K (26.1%) | exon21:c.A3140T;p.H1047L (40%) | exon10:c.G1633A:p.E545K (25%)<br>exon10:c.A1634G:p.E545G (25%) |

ER, estrogen receptor, PgR, progesterone receptor; HER2, human epidermal growth factor receptor 2; FISH, fluorescence in situ hybridization; VAF, variant allele frequency; n.a., not available

